# SARS-CoV-2 antibody prevalence among industrial livestock operation workers and nearby community residents, North Carolina, USA, 2021-2022

**DOI:** 10.1101/2022.10.31.22281764

**Authors:** Carolyn Gigot, Nora Pisanic, Kate Kruczynski, Magdielis Gregory Rivera, Kristoffer Spicer, Kathleen M. Kurowski, Pranay Randad, Kirsten Koehler, William A. Clarke, Phyla Holmes, DJ Hall, Devon Hall, Christopher D. Heaney

## Abstract

Industrial livestock operations (ILOs), particularly processing facilities, emerged as centers of coronavirus disease 2019 (COVID-19) outbreaks in spring 2020. Confirmed cases of COVID-19 underestimate true prevalence. To investigate prevalence of antibodies against SARS-CoV-2, we enrolled 279 participants in North Carolina from February 2021 to July 2022: 90 from households with at least one ILO worker (ILO), 97 from high-ILO intensity areas (ILO neighbors – ILON), and 92 from metropolitan areas (Metro). Participants provided a saliva swab we analyzed for SARS-CoV-2 IgG using a multiplex immunoassay. Prevalence of infection-induced IgG (positive for nucleocapsid and receptor binding domain) was higher among ILO (63%) compared to ILON (42.9%) and Metro (48.7%) participants (prevalence ratio [PR] =1.38; 95% confidence interval [CI]: 1.06, 1.80; ref. ILON and Metro combined). Prevalence of infection-induced IgG was also higher among ILO participants compared to an Atlanta healthcare worker cohort (PR=2.45, 95% CI: 1.8, 3.3) and a general population cohort in North Carolina (PRs 6.37-10.67). Infection-induced IgG prevalence increased over the study period. Participants reporting not masking in public in the past two weeks had higher infection-induced IgG prevalence (78.6%) compared to participants reporting masking (49.3%) (PR=1.59; 95% CI: 1.19, 2.13). Lower education, more people per bedroom, Hispanic/Latino ethnicity, and more contact with people outside the home were also associated with higher infection-induced IgG prevalence. Similar proportions of ILO (51.6%), ILON (48.4%), and Metro (55.4%) participants completed the COVID-19 primary vaccination series; median completion was more than four months later for ILO compared to ILON and Metro participants.

**Importance:** Few studies have measured COVID-19 seroprevalence in North Carolina, especially among rural, Black, and Hispanic/Latino communities that have been heavily affected. Antibody results show high rates of COVID-19 among industrial livestock operation workers and their household members. Antibody results add to evidence of health disparities in COVID-19 by socioeconomic status and ethnicity. Associations between masking and physical distancing with antibody results also add to evidence of the effectiveness of these prevention strategies. Delays in the timing of receipt of COVID-19 vaccination reinforce the importance of dismantling vaccination barriers, especially for industrial livestock operation workers and their household members.

## INTRODUCTION

North Carolina is the second largest hog, third largest turkey, and fourth largest broiler chicken producing state (1). Animal slaughtering and processing workers have more than twice the rate of injury and illness (6.7 per 100 full-time equivalents) and animal production workers close to twice the rate of injury and illness (5.2 per 100 full-time equivalents) compared to all US workers (2.9 per 100 full-time equivalents), despite reporting exemptions and other factors likely resulting in injury and illness undercounts (2–5). Hog and poultry production have also been associated with a range of adverse health outcomes among nearby community residents, including respiratory health problems and infectious diseases (6–8). Since winter 2019, COVID-19 has become a health hazard associated with working at or living near industrial livestock operations (ILOs). Tens of thousands of cases of COVID-19 have been associated with working at meat and poultry processing facilities (9–11). Taylor et al. found an association between county livestock processing plants and county COVID-19 incidence, estimating as many as 8% of US cases through summer 2020 could be linked to processing plants (12). High numbers of COVID-19 cases have also been associated with food processing, food manufacturing, and agricultural workplaces more broadly (13). COVID-19 has disproportionately burdened low income communities of color (14–16). Disparities by race and ethnicity are also evident among livestock processing and agricultural workers (10, 13).

Given limited access to SARS-CoV-2 molecular testing, particularly for asymptomatic or mild COVID-19 cases, and limitations in molecular test reporting, SARS-CoV-2 antibody testing represents an attractive strategy to estimate COVID-19 prevalence, attack rates, and population immunity due to prior infection and/or vaccination (17, 18). SARS-CoV-2 antibodies in oral fluid (hereafter, salivary) have been shown to correspond to SARS-CoV-2 antibodies present in blood and to differentiate between PCR-confirmed cases and pre-COVID-19 samples with high sensitivity and specificity (19–21). Compared to blood collection, saliva collection is painless, safe, and readily self-collected at home and mailed to a testing lab. However, saliva has been less frequently used for surveillance (20, 22).

We used a salivary multiplex immunoassay targeting IgG responses to SARS-CoV-2 nucleocapsid (N), receptor-binding domain (RBD), and spike (S) protein to differentiate between infection-induced versus infection- and/or vaccination-induced immune response. Infection induces antibodies against all proteins, while the mRNA (Pfizer-BioNTech, Moderna), Janssen (Johnson & Johnson), and Novavax vaccines currently approved for use in the United States induce RBD- and S-specific antibodies only (23). Accordingly, individuals testing positive for both SARS-CoV-2 N and RBD IgG likely experienced infection (24).

In this study, we measured salivary SARS-CoV-2 IgG prevalence in a cohort of North Carolina households enrolled in collaboration with REACH (Rural Empowerment Association for Community Help), a community group based in Duplin County, North Carolina. We aimed to (a) compare infection-induced IgG prevalence between participants living in households with at least one adult working at an industrial hog or poultry operation, meatpacking plant, or animal rendering plant (industrial livestock operation household group – ILO), participants living nearby these facilities without any known occupational exposure to livestock (ILO neighbors – ILON), and participants living in metropolitan areas of North Carolina (Metro); (b) identify risk factors for infection-induced IgG prevalence within our study population; and (c) compare infection-induced IgG prevalence between ILO participants and a cohort of other high-risk occupation workers sampled using the same assay, as well as a general population-representative cohort in North Carolina.

## RESULTS

### Participant characteristics

A total of 279 individuals from 240 households (80 ILO, 80 ILON, and 80 Metro) participated (Table 1). ILON participants were generally enrolled earliest, with a median interview date of June 6, 2021, followed by Metro participants (median August 18, 2021), and ILO participants (median February 9, 2022). Overall, ILON participants were older than ILO and Metro participants. Roughly half of ILO and ILON participants were female, compared to 72.8% of Metro participants. Most participants in all groups were Black; more Metro participants were White and fewer were Hispanic/Latino compared to ILO and ILON participants. Education level differed between groups: most ILO participants had a high school education or lower, while most Metro participants had post-high school education. Most participants lived in homes with one or fewer household members per bedroom, although ILO participants reported more household members per bedroom compared to ILON and Metro participants. The majority of participants reported that their household’s primary health care provider was a private doctor or clinic. However, more ILON (15.5%) compared to ILO (13.3%) and Metro (8.7%) participants reported not having health insurance. More than twice as many ILO participants (91.1%) reported working in person compared to ILON (42.3%) and Metro (40.2%) participants.

**Table 1.**
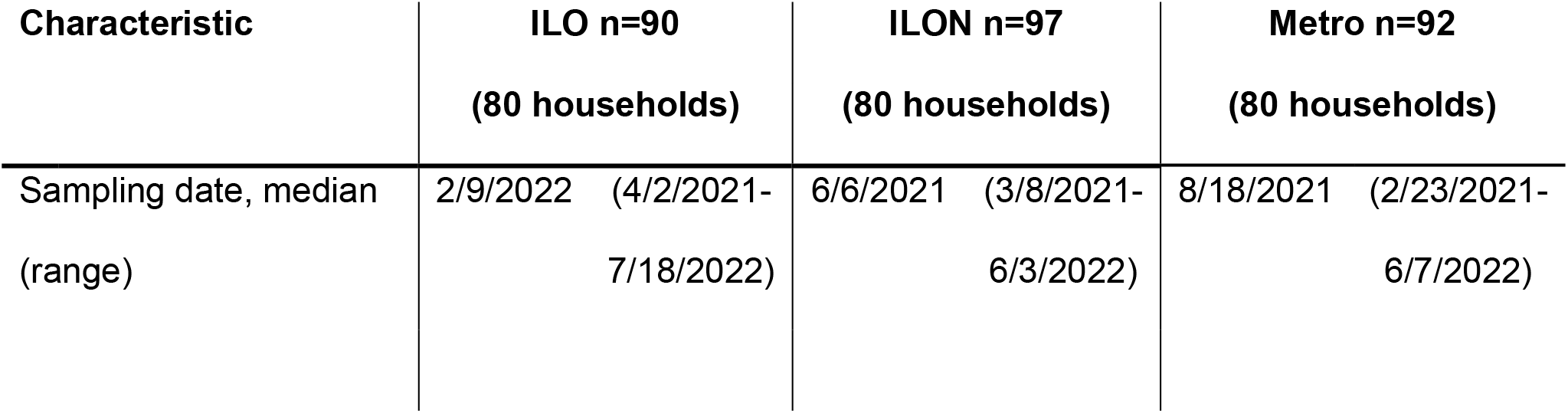
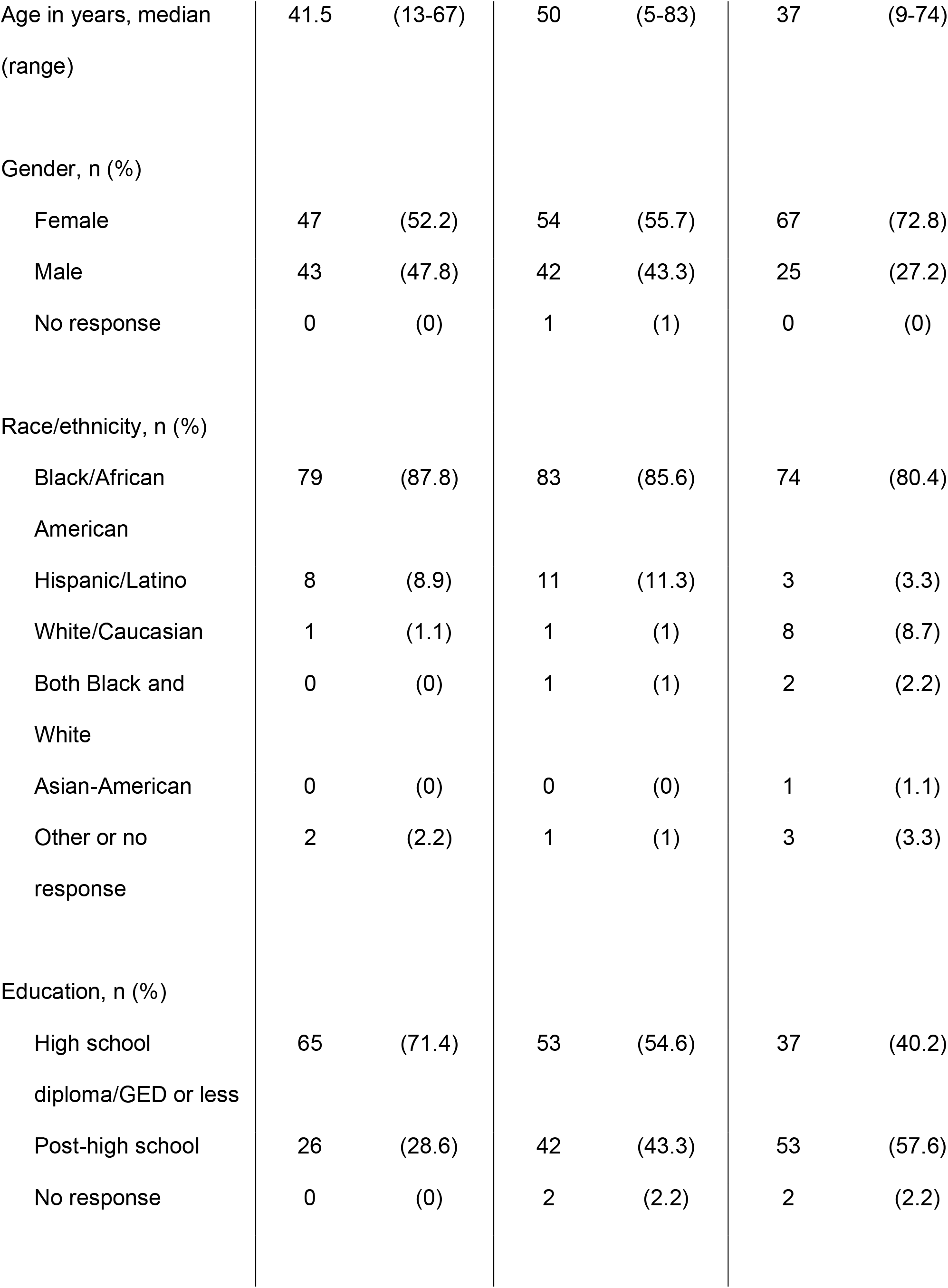
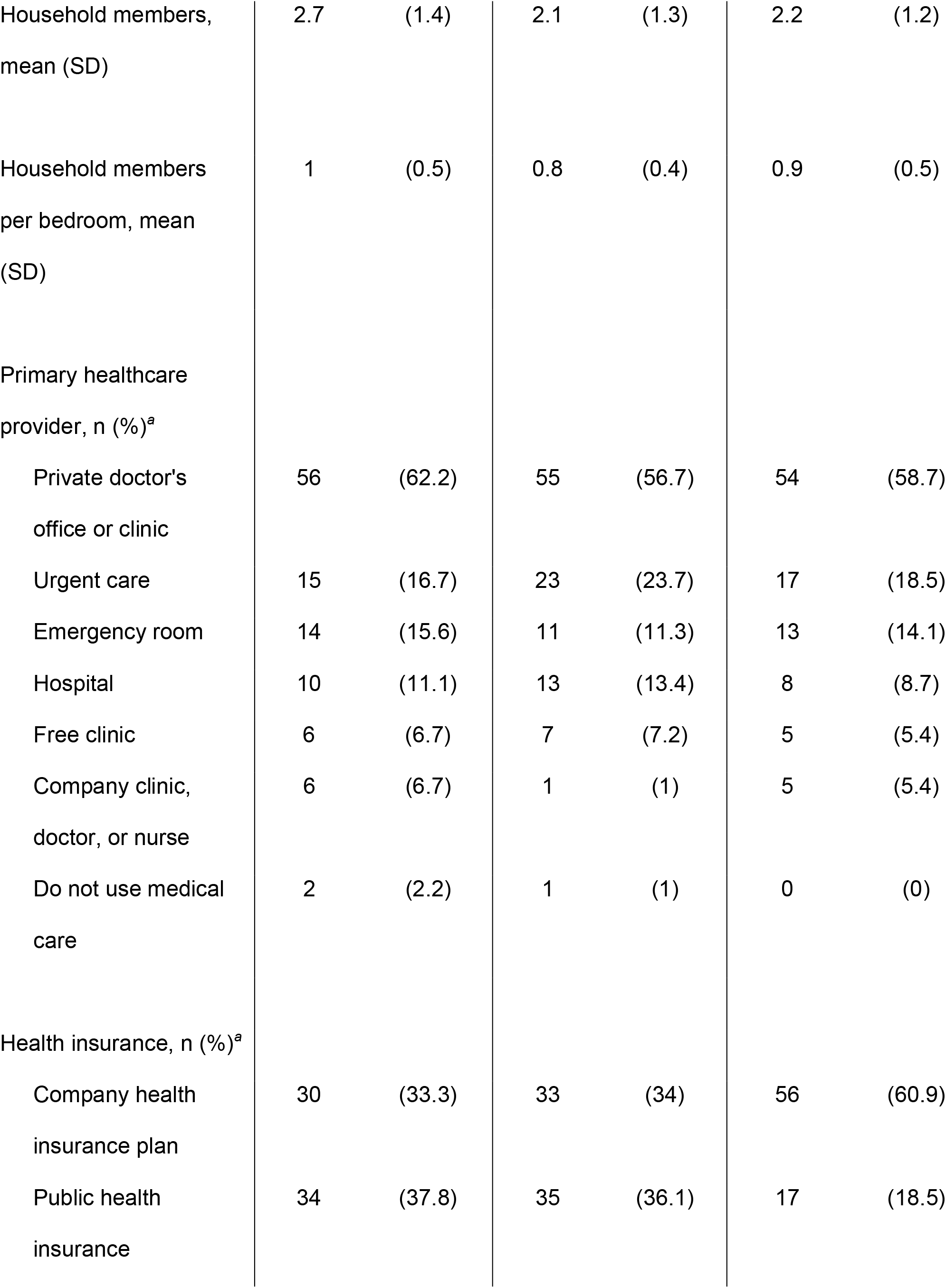
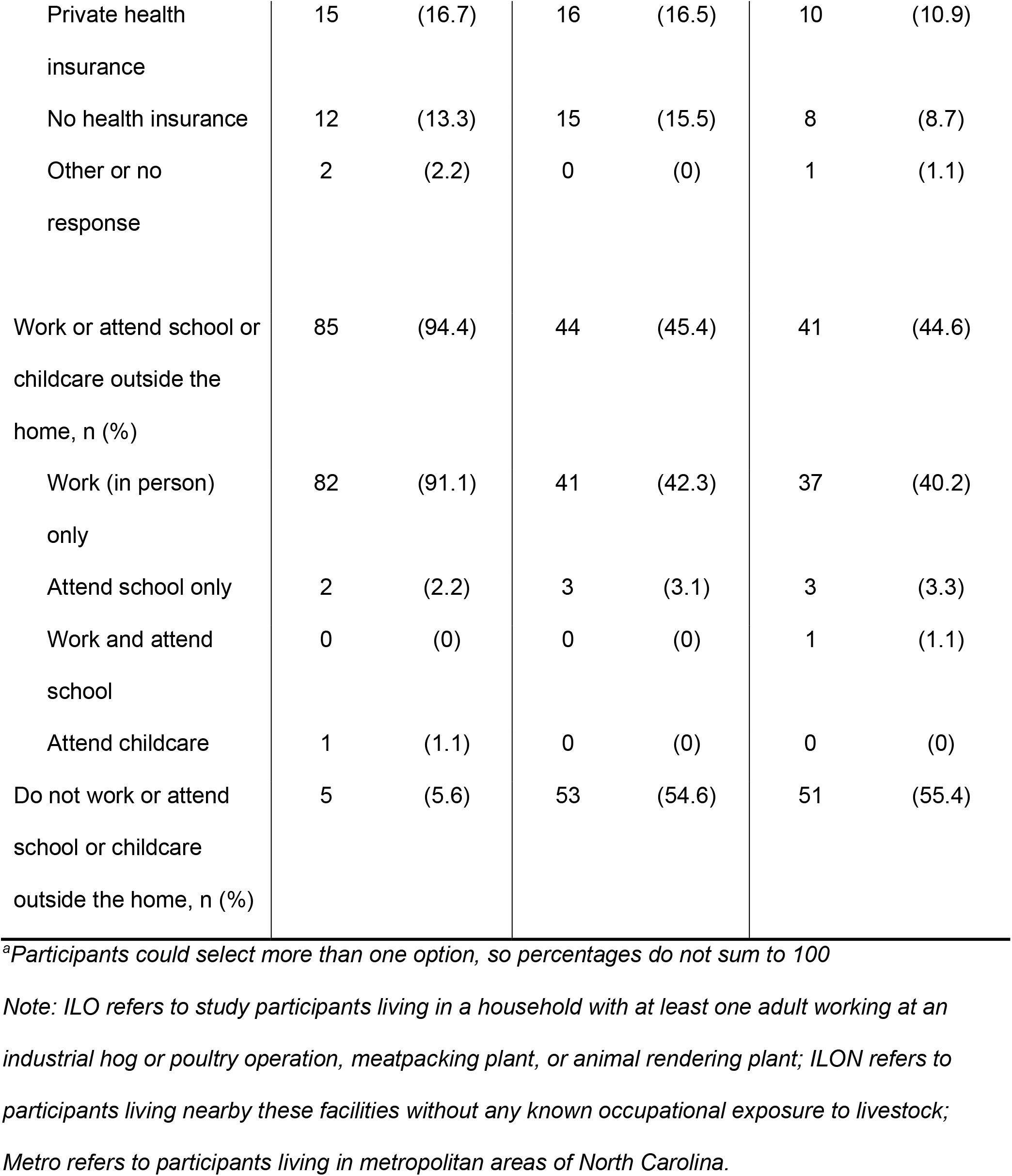
Participant characteristics by study group

| Characteristic | ILO n=90<br>(80 households) |  | ILON n=97<br>(80 households) |  | Metro n=92<br>(80 households) |  |
| --- | --- | --- | --- | --- | --- | --- |
| Sampling date, median<br>(range) | 2/9/2022 | (4/2/2021-7/18/2022) | 6/6/2021 | (3/8/2021-6/3/2022) | 8/18/2021 | (2/23/2021-6/7/2022) |

|  |  |  |  |
| --- | --- | --- | --- |
| Age in years, median<br>(range) | 41.5<br>(13-67) | 50<br>(5-83) | 37<br>(9-74) |
| Gender, n (%) |  |  |  |
| Female | 47<br>(52.2) | 54<br>(55.7) | 67<br>(72.8) |
| Male | 43<br>(47.8) | 42<br>(43.3) | 25<br>(27.2) |
| No response | 0<br>(0) | 1<br>(1) | 0<br>(0) |
| Race/ethnicity, n (%) |  |  |  |
| Black/African<br>American | 79<br>(87.8) | 83<br>(85.6) | 74<br>(80.4) |
| Hispanic/Latino | 8<br>(8.9) | 11<br>(11.3) | 3<br>(3.3) |
| White/Caucasian | 1<br>(1.1) | 1<br>(1) | 8<br>(8.7) |
| Both Black and<br>White | 0<br>(0) | 1<br>(1) | 2<br>(2.2) |
| Asian-American | 0<br>(0) | 0<br>(0) | 1<br>(1.1) |
| Other or no<br>response | 2<br>(2.2) | 1<br>(1) | 3<br>(3.3) |
| Education, n (%) |  |  |  |
| High school<br>diploma/GED or less | 65<br>(71.4) | 53<br>(54.6) | 37<br>(40.2) |
| Post-high school | 26<br>(28.6) | 42<br>(43.3) | 53<br>(57.6) |
| No response | 0<br>(0) | 2<br>(2.2) | 2<br>(2.2) |
| Household members,<br>mean (SD) | 2.7 | (1.4) | 2.1 | (1.3) | 2.2 | (1.2) |
| Household members<br>per bedroom, mean<br>(SD) | 1 | (0.5) | 0.8 | (0.4) | 0.9 | (0.5) |
| Primary healthcare<br>provider, n (%)<sup>a</sup> |  |  |  |  |  |  |
| Private doctor's<br>office or clinic | 56 | (62.2) | 55 | (56.7) | 54 | (58.7) |
| Urgent care | 15 | (16.7) | 23 | (23.7) | 17 | (18.5) |
| Emergency room | 14 | (15.6) | 11 | (11.3) | 13 | (14.1) |
| Hospital | 10 | (11.1) | 13 | (13.4) | 8 | (8.7) |
| Free clinic | 6 | (6.7) | 7 | (7.2) | 5 | (5.4) |
| Company clinic,<br>doctor, or nurse | 6 | (6.7) | 1 | (1) | 5 | (5.4) |
| Do not use medical<br>care | 2 | (2.2) | 1 | (1) | 0 | (0) |
| Health insurance, n (%)<sup>a</sup> |  |  |  |  |  |  |
| Company health<br>insurance plan | 30 | (33.3) | 33 | (34) | 56 | (60.9) |
| Public health<br>insurance | 34 | (37.8) | 35 | (36.1) | 17 | (18.5) |
| Private health insurance | 15 | (16.7) | 16 | (16.5) | 10 | (10.9) |
| No health insurance | 12 | (13.3) | 15 | (15.5) | 8 | (8.7) |
| Other or no response | 2 | (2.2) | 0 | (0) | 1 | (1.1) |
| Work or attend school or childcare outside the home, n (%) | 85 | (94.4) | 44 | (45.4) | 41 | (44.6) |
| Work (in person) only | 82 | (91.1) | 41 | (42.3) | 37 | (40.2) |
| Attend school only | 2 | (2.2) | 3 | (3.1) | 3 | (3.3) |
| Work and attend school | 0 | (0) | 0 | (0) | 1 | (1.1) |
| Attend childcare | 1 | (1.1) | 0 | (0) | 0 | (0) |
| Do not work or attend school or childcare outside the home, n (%) | 5 | (5.6) | 53 | (54.6) | 51 | (55.4) |

<sup>a</sup>Participants could select more than one option, so percentages do not sum to 100
Note: ILO refers to study participants living in a household with at least one adult working at an industrial hog or poultry operation, meatpacking plant, or animal rendering plant; ILON refers to participants living nearby these facilities without any known occupational exposure to livestock; Metro refers to participants living in metropolitan areas of North Carolina.

### COVID-19 vaccination over time by study group

More Metro participants (20.7%) were up to date with COVID-19 vaccines, receiving all doses in the primary series and at least one booster, compared to ILO (13.2%) or ILON (10.3%) participants (Table S1). More Metro participants (55.4%) also completed the COVID-19 primary vaccination series, receiving either a first dose and second dose of the Pfizer, Moderna, or Novavax vaccines or a single dose of the Johnson & Johnson vaccine, compared to ILO (51.6%) or ILON (48.4%) participants (Table S1). There was no statistically significant difference in time to primary series completion or becoming up to date between the study groups (p-values for 3-group log-rank test 0.4 and 0.1 respectively). However, among participants who received a booster dose, the median date of receiving that booster dose was later for ILO (December 19, 2021) compared to ILON (November 3, 2021) and Metro (November 3, 2021) participants, and the same pattern held for primary series completion (Figure 1).

**Figure 1.**
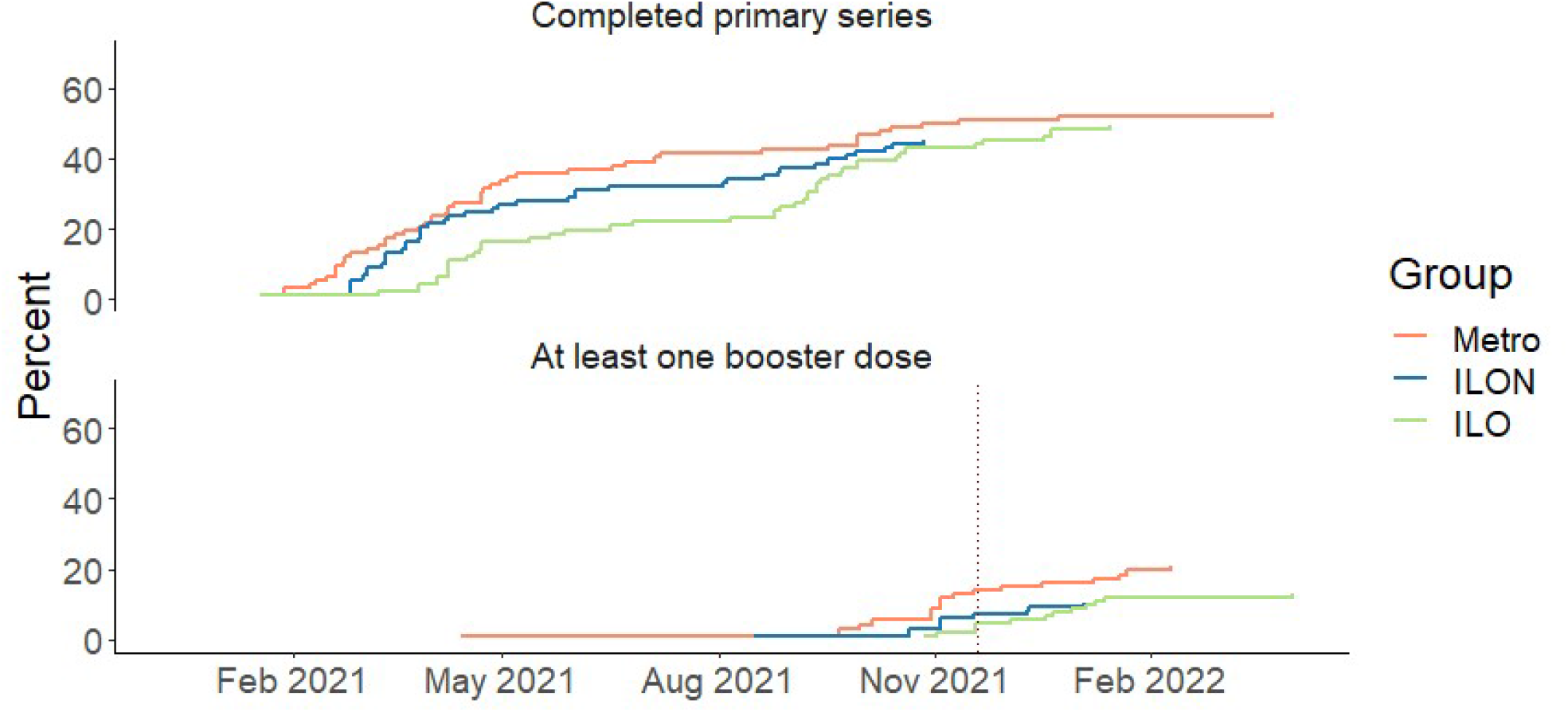
Vaccination over time by study group. Origin is the date of the first FDA Emergency Use Authorization for a vaccine against COVID-19 (Pfizer-BioNTech), December 11, 2020, and dotted line is the date on which CDC expanded eligibility for a booster shot to all adults, November 19, 2021 (25). *Note: ILO refers to study participants living in a household with at least one adult working at an industrial hog or poultry operation, meatpacking plant, or animal rendering plant; ILON refers to participants living nearby these facilities without any known occupational exposure to livestock; Metro refers to participants living in metropolitan areas of North Carolina*.

### SARS-CoV-2 IgG antibody and self-reported COVID-19 outcomes

Most participant saliva samples tested had SARS-CoV-2 IgG antibodies (Table 2). The prevalence of infection-induced IgG (positive for both N and RBD) was higher among ILO (63%) than among ILON (42.9%) and Metro (48.7%) participants (PR=1.38; 95% CI: 1.06, 1.8). The prevalence of infection- and/or vaccination-induced IgG (positive for RBD) was similar among ILO (78.1%), ILON (63.1%), and Metro participants (77.6%). Significantly more ILO participants reported at least one and at least two COVID-19 symptoms compared to ILON and Metro participants (26). Fewer participants reported thinking they had COVID-19 than had infection-induced IgG. Even fewer participants reported they had ever tested positive for SARS-CoV-2, with the highest proportion among Metro participants (21.1%), followed by ILON (11.9%) and ILO (13.7%) participants.

**Table 2.**
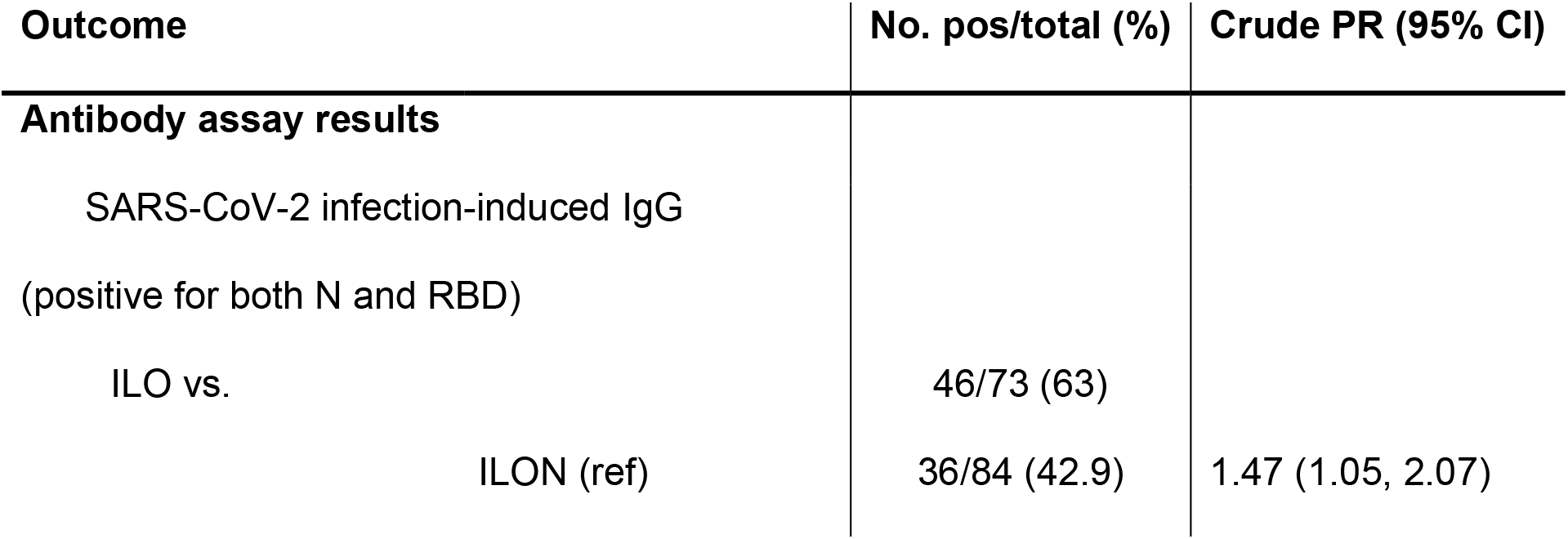
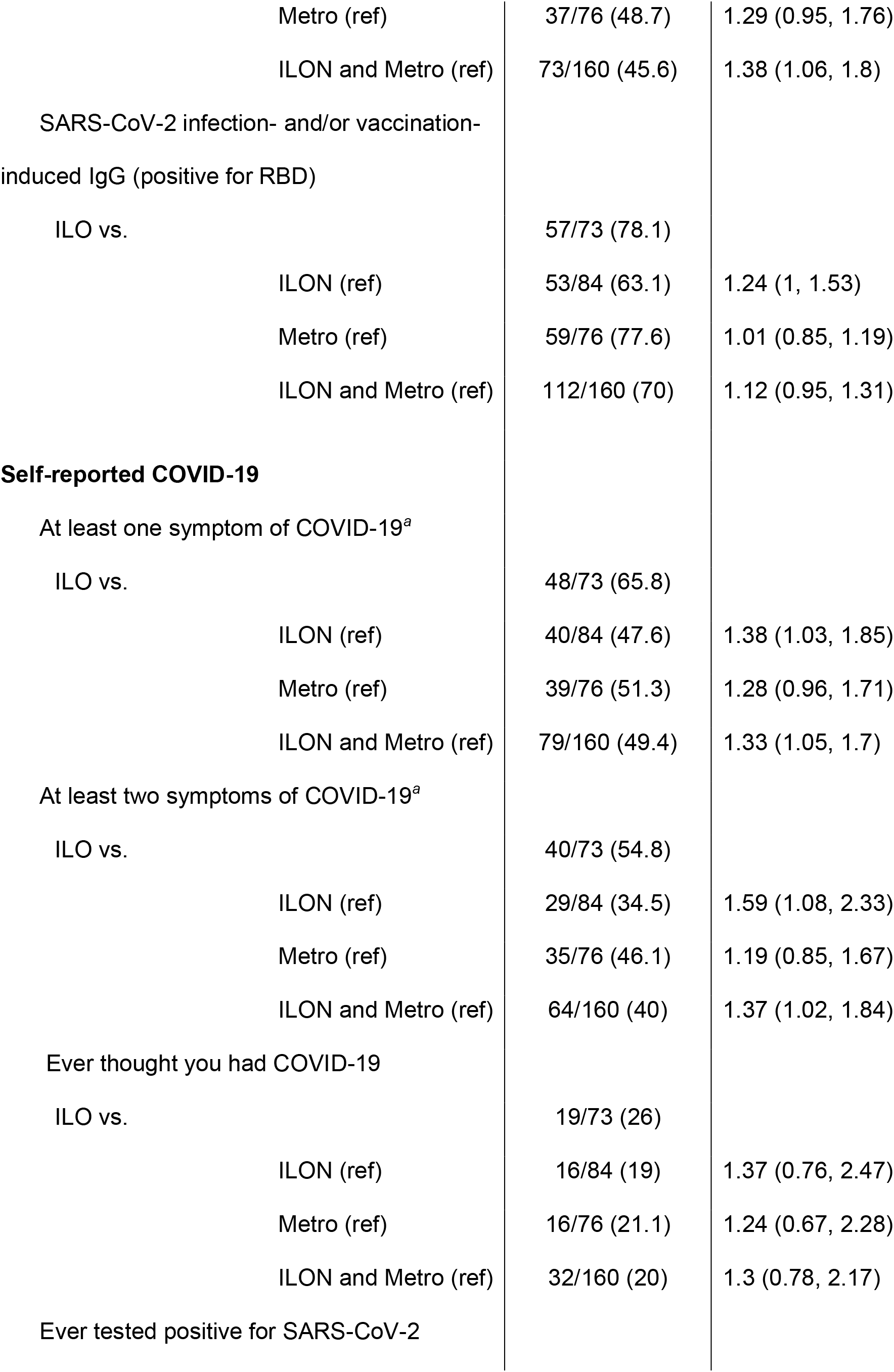
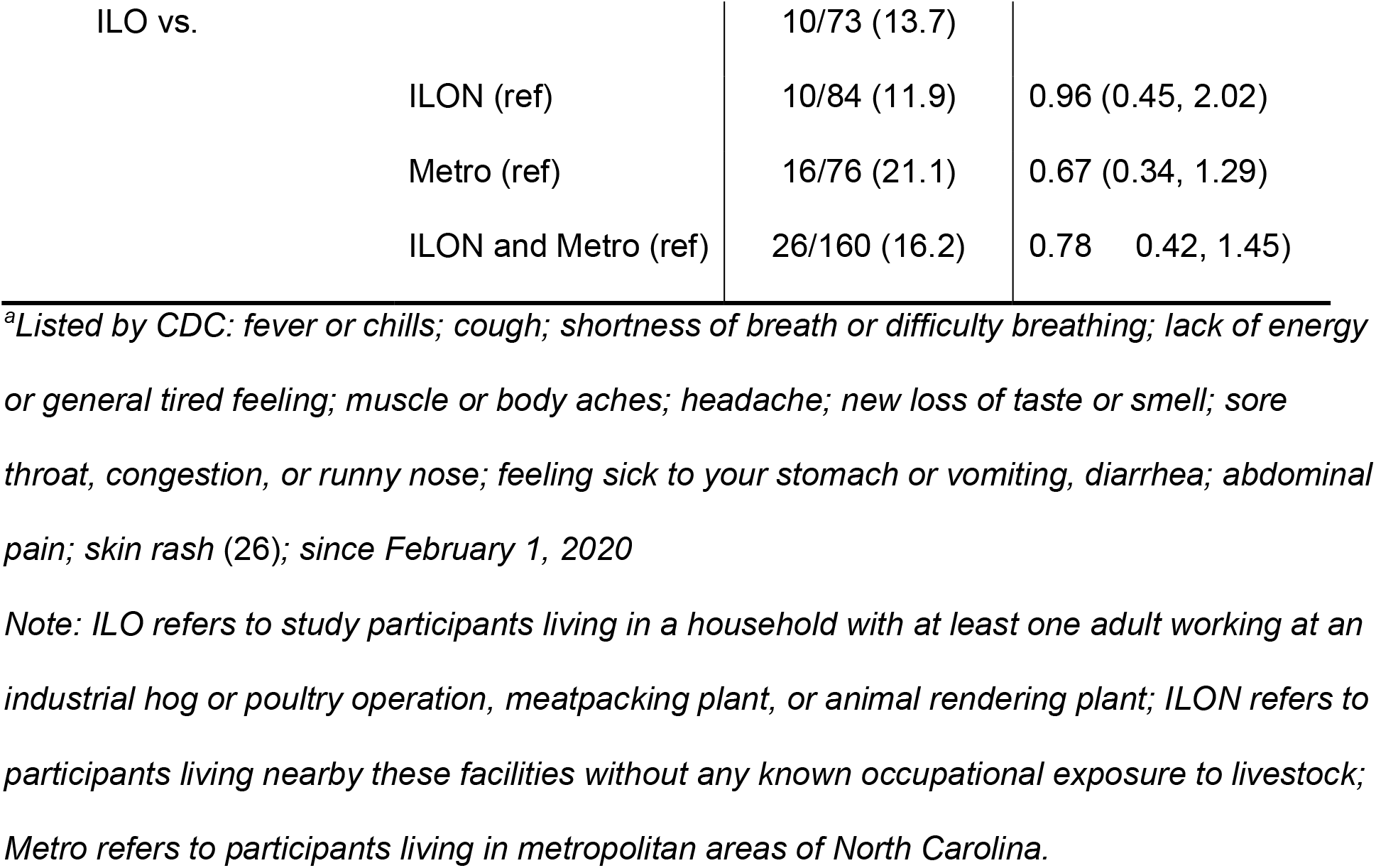
SARS-CoV-2 antibody assay results and self-reported COVID-19 prevalence and prevalence ratios by study group

| Outcome | No. pos/total (%) | Crude PR (95% CI) |
| --- | --- | --- |
| **Antibody assay results** |  |  |
| SARS-CoV-2 infection-induced IgG<br>(positive for both N and RBD) |  |  |
| ILO vs. | 46/73 (63) |  |
| ILON (ref) | 36/84 (42.9) | 1.47 (1.05, 2.07) |

|  |  |  |  |
| --- | --- | --- | --- |
|  | Metro (ref) | 37/76 (48.7) | 1.29 (0.95, 1.76) |
|  | ILON and Metro (ref) | 73/160 (45.6) | 1.38 (1.06, 1.8) |
| SARS-CoV-2 infection- and/or vaccination- |  |  |  |
| induced IgG (positive for RBD) |  |  |  |
| ILO vs. |  | 57/73 (78.1) |  |
|  | ILON (ref) | 53/84 (63.1) | 1.24 (1, 1.53) |
|  | Metro (ref) | 59/76 (77.6) | 1.01 (0.85, 1.19) |
|  | ILON and Metro (ref) | 112/160 (70) | 1.12 (0.95, 1.31) |
| **Self-reported COVID-19** |  |  |  |
| At least one symptom of COVID-19<sup>a</sup> |  |  |  |
| ILO vs. |  | 48/73 (65.8) |  |
|  | ILON (ref) | 40/84 (47.6) | 1.38 (1.03, 1.85) |
|  | Metro (ref) | 39/76 (51.3) | 1.28 (0.96, 1.71) |
|  | ILON and Metro (ref) | 79/160 (49.4) | 1.33 (1.05, 1.7) |
| At least two symptoms of COVID-19<sup>a</sup> |  |  |  |
| ILO vs. |  | 40/73 (54.8) |  |
|  | ILON (ref) | 29/84 (34.5) | 1.59 (1.08, 2.33) |
|  | Metro (ref) | 35/76 (46.1) | 1.19 (0.85, 1.67) |
|  | ILON and Metro (ref) | 64/160 (40) | 1.37 (1.02, 1.84) |
| Ever thought you had COVID-19 |  |  |  |
| ILO vs. |  | 19/73 (26) |  |
|  | ILON (ref) | 16/84 (19) | 1.37 (0.76, 2.47) |
|  | Metro (ref) | 16/76 (21.1) | 1.24 (0.67, 2.28) |
|  | ILON and Metro (ref) | 32/160 (20) | 1.3 (0.78, 2.17) |
| Ever tested positive for SARS-CoV-2 |  |  |  |
| ILO vs. |  | 10/73 (13.7) |  |
|  | ILON (ref) | 10/84 (11.9) | 0.96 (0.45, 2.02) |
|  | Metro (ref) | 16/76 (21.1) | 0.67 (0.34, 1.29) |
|  | ILON and Metro (ref) | 26/160 (16.2) | 0.78 0.42, 1.45) |

<sup>a</sup>Listed by CDC: fever or chills; cough; shortness of breath or difficulty breathing; lack of energy or general tired feeling; muscle or body aches; headache; new loss of taste or smell; sore throat, congestion, or runny nose; feeling sick to your stomach or vomiting, diarrhea; abdominal pain; skin rash (26); since February 1, 2020
Note: ILO refers to study participants living in a household with at least one adult working at an industrial hog or poultry operation, meatpacking plant, or animal rendering plant; ILON refers to participants living nearby these facilities without any known occupational exposure to livestock; Metro refers to participants living in metropolitan areas of North Carolina.

### SARS-CoV-2 infection-induced IgG prevalence by participant characteristics

The proportion of participants with salivary SARS-CoV-2 infection-induced IgG increased over the study period (Table 3). Several participant demographic characteristics and infection prevention behaviors were associated with infection-induced IgG prevalence. The strongest association was for participants who reported generally wearing a mask in public in the past two weeks. Participants who reported not wearing a mask had significantly higher infection-induced IgG prevalence (78.6%) compared to participants who reported wearing a mask (49.3%) (PR=1.59; 95% CI: 1.19, 2.13). Participants with greater than a high school education had significantly lower infection-induced IgG prevalence (38.1%) compared to participants with a high school education or less (60.2%) (PR=0.63, 95% CI:0.48, 0.84). Participants who lived in households with more than one person per bedroom had significantly higher infection-induced IgG prevalence (69.4%) compared to participants in households with one person or fewer per bedroom (46.4%) (PR=1.5, 95% CI:1.15, 1.95). Hispanic/Latino participants had significantly higher infection-induced IgG prevalence (72.7%) compared to Black participants (49.7%) (PR=1.46; 95% CI: 1.1, 1.94). Only 16.7% of White participants had infection-induced IgG. Reduced contact with people outside the home was associated with reduced infection-induced IgG prevalence.

**Table 3.**
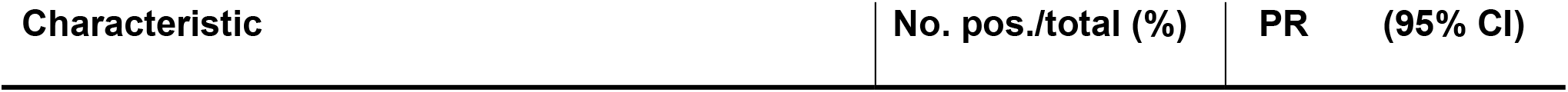
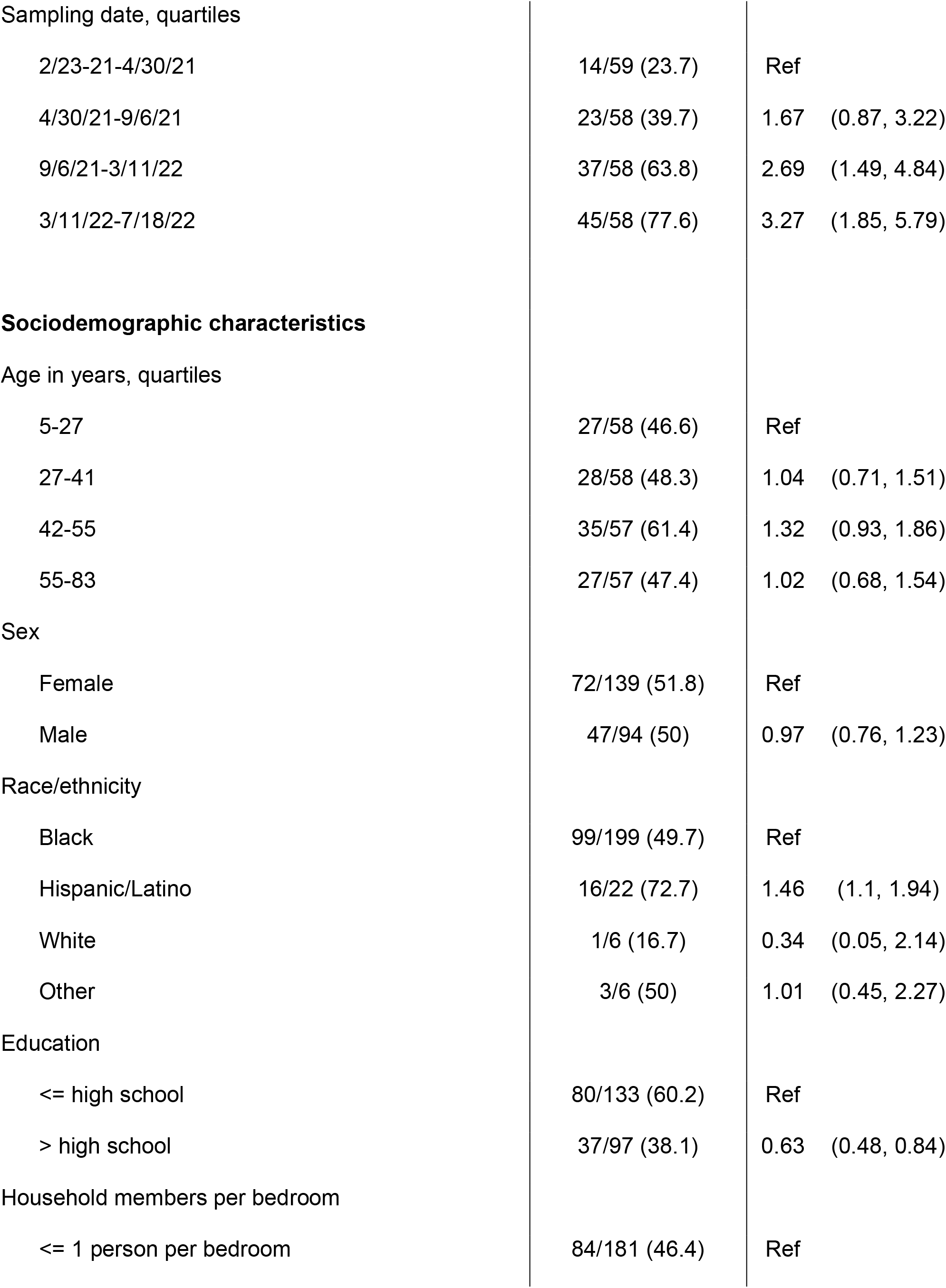
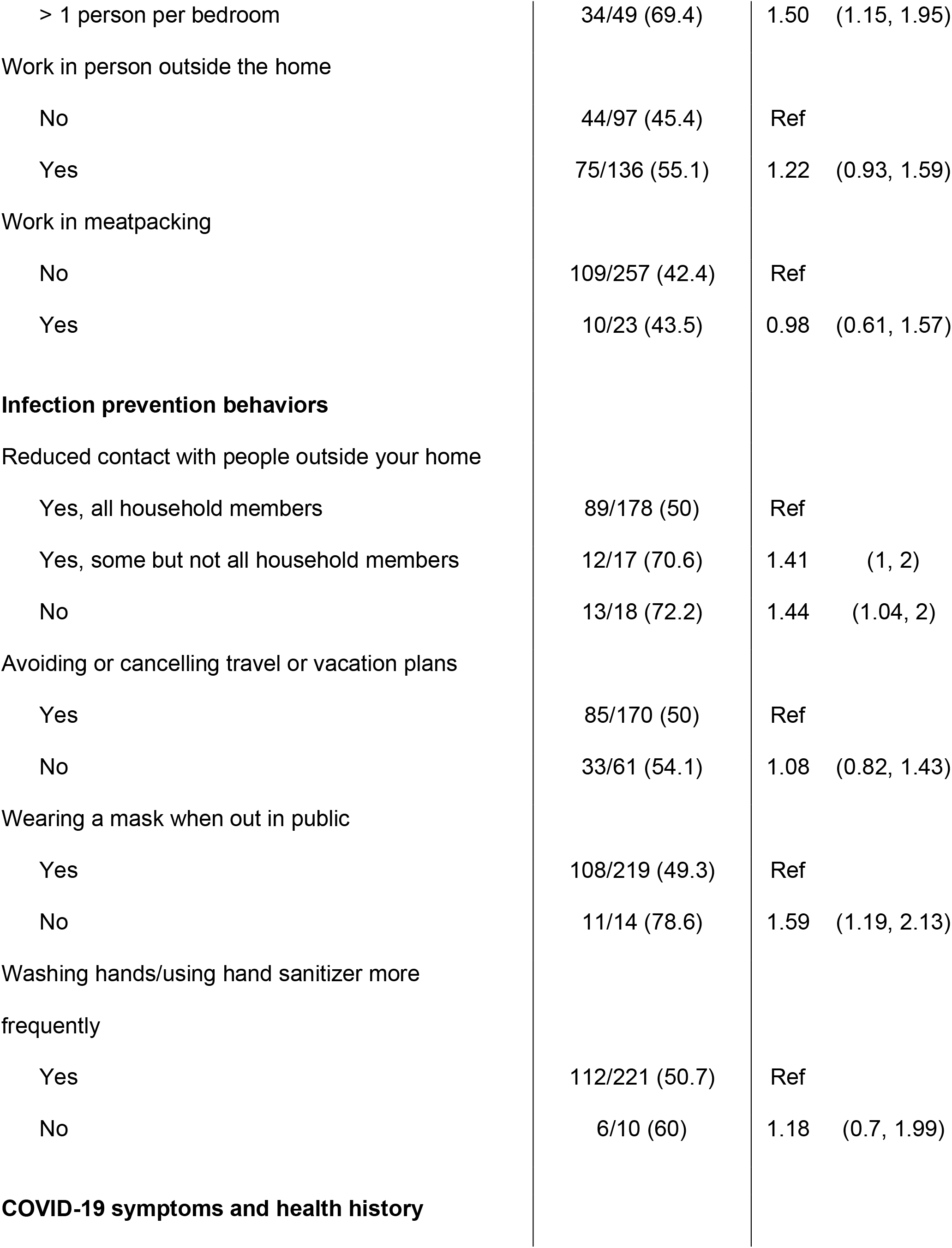
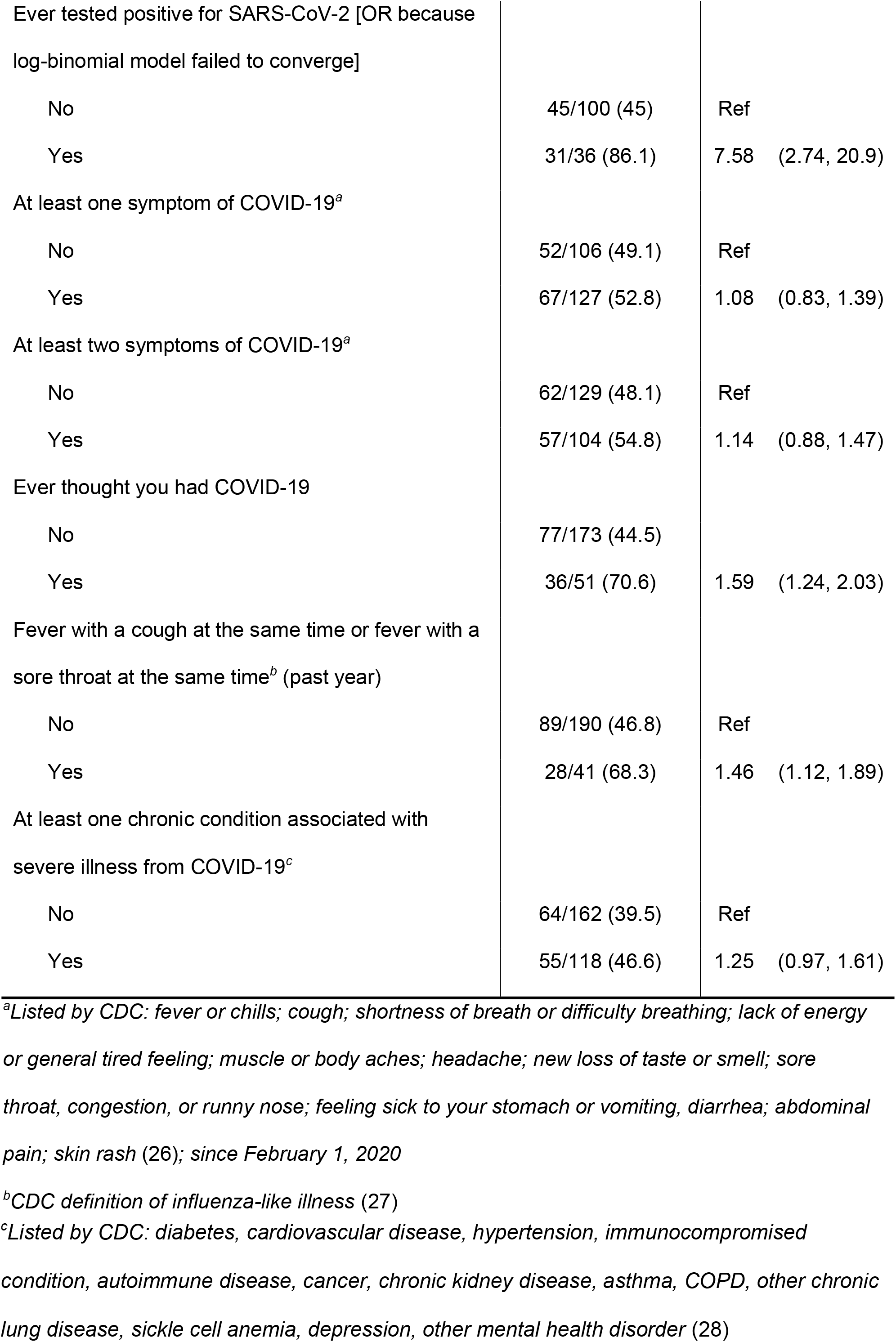
SARS-CoV-2 infection-induced IgG prevalence and prevalence ratios (PR) and 95% confidence intervals (CI) by participant characteristics, North Carolina, USA, 2021-2022.

Sampling date, quartiles
|  |  |  |  |
| --- | --- | --- | --- |
| 2/23-21-4/30/21 | 14/59 (23.7) | Ref |  |
| 4/30/21-9/6/21 | 23/58 (39.7) | 1.67 | (0.87, 3.22) |
| 9/6/21-3/11/22 | 37/58 (63.8) | 2.69 | (1.49, 4.84) |
| 3/11/22-7/18/22 | 45/58 (77.6) | 3.27 | (1.85, 5.79) |
| Age in years, quartiles |  |  |  |
| 5-27 | 27/58 (46.6) | Ref |  |
| 27-41 | 28/58 (48.3) | 1.04 | (0.71, 1.51) |
| 42-55 | 35/57 (61.4) | 1.32 | (0.93, 1.86) |
| 55-83 | 27/57 (47.4) | 1.02 | (0.68, 1.54) |
| Sex |  |  |  |
| Female | 72/139 (51.8) | Ref |  |
| Male | 47/94 (50) | 0.97 | (0.76, 1.23) |
| Race/ethnicity |  |  |  |
| Black | 99/199 (49.7) | Ref |  |
| Hispanic/Latino | 16/22 (72.7) | 1.46 | (1.1, 1.94) |
| White | 1/6 (16.7) | 0.34 | (0.05, 2.14) |
| Other | 3/6 (50) | 1.01 | (0.45, 2.27) |
| Education |  |  |  |
| <= high school | 80/133 (60.2) | Ref |  |
| > high school | 37/97 (38.1) | 0.63 | (0.48, 0.84) |
| Household members per bedroom |  |  |  |
| <= 1 person per bedroom | 84/181 (46.4) | Ref |  |
| > 1 person per bedroom | 34/49 (69.4) | 1.50 | (1.15, 1.95) |
| Work in person outside the home |  |  |  |
| No | 44/97 (45.4) | Ref |  |
| Yes | 75/136 (55.1) | 1.22 | (0.93, 1.59) |
| Work in meatpacking |  |  |  |
| No | 109/257 (42.4) | Ref |  |
| Yes | 10/23 (43.5) | 0.98 | (0.61, 1.57) |
| **Infection prevention behaviors** |  |  |  |
| Reduced contact with people outside your home |  |  |  |
| Yes, all household members | 89/178 (50) | Ref |  |
| Yes, some but not all household members | 12/17 (70.6) | 1.41 | (1, 2) |
| No | 13/18 (72.2) | 1.44 | (1.04, 2) |
| Avoiding or cancelling travel or vacation plans |  |  |  |
| Yes | 85/170 (50) | Ref |  |
| No | 33/61 (54.1) | 1.08 | (0.82, 1.43) |
| Wearing a mask when out in public |  |  |  |
| Yes | 108/219 (49.3) | Ref |  |
| No | 11/14 (78.6) | 1.59 | (1.19, 2.13) |
| Washing hands/using hand sanitizer more frequently |  |  |  |
| Yes | 112/221 (50.7) | Ref |  |
| No | 6/10 (60) | 1.18 | (0.7, 1.99) |
| **COVID-19 symptoms and health history** |  |  |  |

|  |  |  |
| --- | --- | --- |
| Ever tested positive for SARS-CoV-2 [OR because log-binomial model failed to converge] |  |  |
| No | 45/100 (45) | Ref |
| Yes | 31/36 (86.1) | 7.58 (2.74, 20.9) |
| At least one symptom of COVID-19<sup>a</sup> |  |  |
| No | 52/106 (49.1) | Ref |
| Yes | 67/127 (52.8) | 1.08 (0.83, 1.39) |
| At least two symptoms of COVID-19<sup>a</sup> |  |  |
| No | 62/129 (48.1) | Ref |
| Yes | 57/104 (54.8) | 1.14 (0.88, 1.47) |
| Ever thought you had COVID-19 |  |  |
| No | 77/173 (44.5) |  |
| Yes | 36/51 (70.6) | 1.59 (1.24, 2.03) |
| Fever with a cough at the same time or fever with a sore throat at the same time<sup>b</sup> (past year) |  |  |
| No | 89/190 (46.8) | Ref |
| Yes | 28/41 (68.3) | 1.46 (1.12, 1.89) |
| At least one chronic condition associated with severe illness from COVID-19<sup>c</sup> |  |  |
| No | 64/162 (39.5) | Ref |
| Yes | 55/118 (46.6) | 1.25 (0.97, 1.61) |

<sup>a</sup>Listed by CDC: fever or chills; cough; shortness of breath or difficulty breathing; lack of energy or general tired feeling; muscle or body aches; headache; new loss of taste or smell; sore throat, congestion, or runny nose; feeling sick to your stomach or vomiting, diarrhea; abdominal pain; skin rash (26); since February 1, 2020
<sup>b</sup>CDC definition of influenza-like illness (27)
<sup>c</sup>Listed by CDC: diabetes, cardiovascular disease, hypertension, immunocompromised condition, autoimmune disease, cancer, chronic kidney disease, asthma, COPD, other chronic lung disease, sickle cell anemia, depression, other mental health disorder (28)

Participants’ COVID-19 symptoms and health history also corresponded with SARS-CoV-2 IgG. Participants who reported ever testing positive for SARS-CoV-2 had significantly higher infection-induced IgG prevalence (86.1%) compared to those who reported never testing positive (45%) (OR 7.58; 95% CI: 2.74, 20.9). While participants’ report of at least one or at least two COVID-19 symptoms listed by the CDC were not associated with infection-induced IgG prevalence, participants who thought they had COVID-19 had higher infection-induced IgG prevalence (70.6%) compared with participants who thought they had not (44.5%) (PR=1.59, 95% CI: 1.24, 2.03). Participants who reported fever plus cough or sore throat had higher infection-induced IgG prevalence (68.3%) compared to those who did not (46.8%) (27) (PR=1.46, 95% CI: 1.12, 1.89). Almost half (46.6%) of participants who reported a chronic medical condition listed by CDC as increasing the risk of getting very sick with COVID-19 had infection-induced IgG (28).

The highest correlation between factors associated with SARS-CoV-2 infection-induced IgG indicative of infection was between date quartile and vaccination status (Cramer’s v-test value 0.42), followed by date and study group (0.39), group and education level (0.25), group and household members per bedroom (0.23), and date and level of contact with people outside the home (0.34) (Figure S1).

### SARS-CoV-2 infection-induced IgG prevalence compared to other southern US cohorts

The prevalence of SARS-CoV-2 infection-induced IgG was significantly higher in the ILO group of our study population compared to two other southern US cohorts sampled at times overlapping the enrollment and interview dates of our cohort. Infection-induced IgG prevalence was significantly higher in the ILO group of our study population sampled between March 2021 and July 2022 (63%) compared to the COVID-19 Prevention in Emory Healthcare Personnel (COPE) Study cohort sampled between January and December 2021 (23.2%) using the same salivary multiplex assay (PR=2.45, 95% CI: 1.80, 3.33) (M. H. Collins and C. D. Heaney, correspondence) (29, 30) (Figure 2). Infection-induced IgG prevalence was also significantly higher in the ILO group of our study population (63%) compared to the MURDOCK Cabarrus County COVID-19 Prevalence and Immunity (C3PI) Study cohort, representative of Cabarrus County, North Carolina, sampled March and monthly June through November 2021, using blood testing with the Abbot Alinity N IgG assay (5.9% to 9.9%; PR range 6.37 to 10.67) (L. K. Newby and D. Wixted, correspondence) (31).

**Figure 2.**
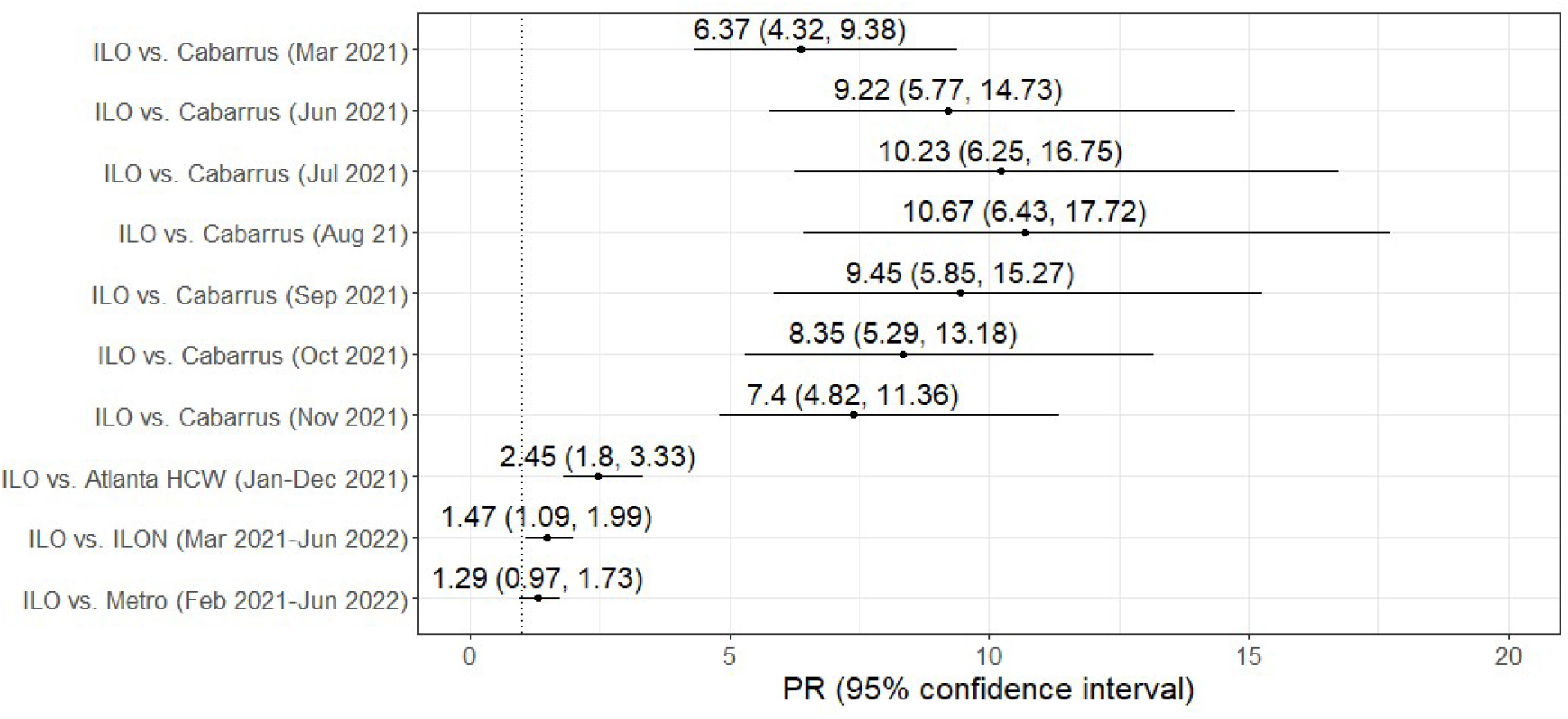
SARS-CoV-2 infection-induced IgG prevalence ratios (PRs) among ILO household group participants (during Apr 2021-Jul 2022) in this study compared to other southern US reference populations. *Note: ILO refers to study participants living in a household with at least one adult working at an industrial hog or poultry operation, meatpacking plant, or animal rendering plant; ILON refers to participants living nearby these facilities without any known occupational exposure to livestock; Metro refers to participants living in metropolitan areas of North Carolina; Cabarrus refers to C3PI Cabarrus County, NC general population-representative cohort; Atlanta HCW refers to Emory COPE health care worker cohort*.

## DISCUSSION

We measured salivary SARS-CoV-2 IgG prevalence among 279 participants, 94% of whom were Black or Hispanic/Latino, underrepresented groups in SARS-CoV-2 seroprevalence surveys. To our knowledge, this study presents the first estimates of SARS-CoV-2 infection-induced antibody prevalence among industrial livestock operation (ILO) workers and their household members, which we observed to be high (63%) compared to participants with no household members working at industrial livestock operation (45.6%) (Table 2). This is consistent with research connecting the agricultural sector and meatpacking facilities with COVID-19 transmission among workers, and connecting meatpacking facilities with transmission in nearby communities (10, 12, 13, 32). However, neither work at a meatpacking facility nor work in person outside the home were associated with elevated prevalence of SARS-CoV-2 infection-induced IgG in our study population (Table 3), and participants living in areas of high industrial livestock operation intensity did not have a higher prevalence (42.9%) compared to metropolitan-area participants (48.7%) (Table 2). This may be due to the relatively small number of meatpacking workers (n=23) and relatively large number of participants with other high-COVID-19-risk jobs and factors (e.g., low income and communities of color). Another contributing factor could be case rate convergence over time between high intensity livestock operation areas and metro areas. As the prevalence of COVID-19 increased in summer and fall 2020, the importance of any single transmission route decreased; also, if many meatpacking workers and nearby residents were infected earlier on, those communities might have a greater rate of at least temporary immunity (32). Our group definitions of ILO, ILON, and Metro also collapse many differences that have been connected to exposure to SARS-CoV-2 and COVID-19: time, age, sex, race/ethnicity, education, and urbanicity, among others (14–16).

Self-reported symptoms of COVID-19 were also higher in the ILO compared to combined ILO and Metro groups (Table 2). This could be because of more COVID-19 cases, though some contribution could also be from other health effects related to industrial livestock production and processing work. Hog and poultry production work have been associated with respiratory and infectious disease broadly, and processing work has also been associated with respiratory, infectious, and skin disease, which overlap with almost all COVID symptoms listed by CDC (6, 26, 33, 34). While 51% of participants overall had SARS-CoV-2 infection-induced IgG, less than half reported thinking they had COVID-19, and only 15.4% reported ever testing positive for COVID-19 (Table 2). Of participants who tested positive for infection-induced IgG, 68% did not think they had COVID-19 (Table 3). This is a higher proportion compared to the estimated prevalence of asymptomatic infections among people with confirmed COVID-19, 40-45% (35). Participants might have attributed any symptoms to other health issues, including some related to ILO work or residence near ILOs. The lower percentage of people ever testing positive for COVID-19 even compared to the low percentage of people who thought they had COVID also underlines the importance of accessible COVID-19 testing.

A higher proportion of Metro compared to ILO or ILON participants completed the primary vaccination series and received at least one booster, although the difference between groups was not statistically significant, and the groups had similar SARS-CoV-2 infection- and/or vaccination-induced IgG (positive for RBD) (Figure 1, Table 2). Among participants who completed the primary vaccination series, the median date of completion was later for ILO compared to ILON and Metro participants, and the same pattern held for booster doses (Figure 1). Although the differences in vaccination timing between groups were not statistically significant, these delays are notable because of the spread of the more-contagious Delta and Omicron variants in summer and winter 2021, respectively (36). Because a greater proportion of ILO participants had a high school education or less, our results are also consistent with evidence of vaccination disparities by social class, and with evidence that vaccination coverage increased most during spring and summer 2021 among people with lower education and income (37).

COVID-19 vaccination rates in our study population were lower than the US and North Carolina general populations. The proportion of our study population (ILO, ILON, and Metro combined) who completed the initial vaccination protocol (52%) was lower than the proportion of North Carolina (62.9%) and US residents (67.2%) vaccinated at the last date of follow-up, July 18, 2022 (38) (Table S1). However, participants may have become vaccinated after their initial or follow-up call; the proportion of participants who had completed the initial vaccination protocol by the median initial or follow-up call date was 50.7%, closer to the proportion of North Carolina (57.3%) and the US (62.6%) at that date, January 10, 2022. The proportion of our cohort who received at least one booster dose was also lower compared to North Carolina and the US at the end of the study, but similar at the median follow-up date (38). Although our modest sample size and timing of initial and follow-up call complicate comparison to North Carolina and the US, our results support the importance of dismantling vaccination barriers, especially for ILO workers, their household members, and rural communities.

The proportion of participants with SARS-CoV-2 infection-induced IgG prevalence increased over the course of the study (Table 3). This is consistent with the spread of the virus over time and trends in other seroprevalence surveys (24, 39). Among health behaviors assessed, wearing a mask had the highest protective effect (Table 3). This is consistent with cohort, ecological, and modelling studies on the efficacy of masks for COVID-19 protection (40). Reporting mask use could also be an indicator of other modifiable and non-modifiable risk factors. We did not ask about the frequency of mask use overall, mask use in particular contexts that might be higher risk transmission settings, or about the type(s) of masks participants used, all of which affect any relationship between mask use and SARS-CoV-2 exposure.

Education level and ethnicity were also associated with SARS-CoV-2 infection-induced IgG. A study of the joint effects of socioeconomic position estimated by education level, race/ethnicity, and gender on COVID-19 mortality among working-age adults found the same trends of higher COVID-19 mortality for low vs. high socioeconomic position adults and for Black and Hispanic/Latino vs. White adults (41). Our results are also consistent with elevated rates of infection-induced seroprevalence among Hispanic/Latino and Black compared with White blood donors across the US (24) and elevated infection-induced seroprevalence among Hispanic/Latino and Black North Carolina residents in surveillance based on hospital remnant blood samples (16), as well as with disproportionate numbers of cases and deaths among Hispanic/Latino, Native American, and Black communities (14). People with fewer socioeconomic resources are less able to use different strategies to avoid exposure to SARS-CoV-2 and more likely to work in crowded occupations or occupations with contact with the public (42). Under racialized capitalism, Hispanic/Latino, Indigenous, and Black workers face occupational status disadvantages even within specific jobs (42, 43).

We found that participants with more than one person per household bedroom had higher prevalence of SARS-CoV-2 infection-induced IgG (Table 3). Living in more crowded conditions could increase exposures to SARS-CoV-2 from household members. Level of contact with people outside of the home was also associated with higher infection-induced IgG prevalence (Table 3). A systematic review of observational studies of SARS-CoV-2 and the betacoronaviruses that cause severe acute respiratory syndrome (SARS-CoV-1) and Middle east respiratory syndrome (MERS) found reduced transmission of viruses with physical distancing of 1m or more as well as with face mask usage, consistent with our results (44).

Participants’ COVID-19 health history and symptoms also corresponded with SARS-CoV-2 IgG prevalence. Close to 90% of participants who reported ever testing positive for SARS-CoV-2 had infection-induced IgG (positive for both N and RBD), compared to 45% of participants who did not report ever testing positive (Table 3). Nucleocapsid (N) IgG is useful for determining response to infection because these antibodies are produced in the immune response to infection and not in the response to vaccines currently approved for use in the US (23). However, a limitation is that N IgG half-life in the body is generally shorter compared to RBD and S IgG (45). Participants who reported testing positive for SARS-CoV-2 but tested negative for N IgG could have been infected longer ago and have levels of N IgG below the positivity cutoff. Participants who reported thinking they had COVID-19 and participants who reported influenza-like illness had a higher prevalence of infection-induced IgG compared to participants who did not, although there was a similar prevalence of infection-induced IgG among participants who reported at least one or at least two symptoms of COVID-19 compared to those who did not (Table 3). This is consistent with relatively high proportions of asymptomatic infections (35), overlap between COVID-19 and other health condition symptoms, and limited durability of N IgG response.

We found prevalence of SARS-CoV-2 infection-induced IgG higher than in comparable cohorts. Infection-induced IgG prevalence in the ILO group of our study population was more than twice that in Emory University’s COPE cohort of Atlanta healthcare workers sampled from January to December 2021 using the same salivary multiplex assay (M. H. Collins and C. D. Heaney, correspondence) (29, 30) (Figure 2). Healthcare workers are at higher risk for COVID-19 compared to the general population (46, 47). High infection-induced IgG among ILO participants compared to workers in another high-COVID-19-risk occupation sampled with the same assay during an overlapping time period underlines the high exposures among North Carolina livestock operation workers and their household members. Infection-induced IgG prevalence among ILO participants was more than five times the highest prevalence observed in March and monthly June through November 2021 in Duke University’s Cabarrus County, North Carolina general-population-representative cohort (L. K. Newby and D. Wixted, correspondence) (31) (Figure 2). Infection-induced IgG prevalence was also higher in the ILO group of our study population (63%) compared to nationwide serology estimates. A study of blood donations estimated infection-induced seroprevalence to be 28.8% overall in December 2021; higher among Hispanic (40.2%) and Black (32.5%) donors and donors living in the South (33.5%) during the same time period (24). Infection-induced IgG prevalence in our cohort was also generally higher compared to estimates using residual data from commercial labs across the US weighted by age, sex, and metropolitan status, which ranged from 20.8% to 57.7% nationally and 22.5% to 52% in North Carolina during the sixteen sampling periods that overlapped our study period (36, 39) (Figure S2).

An important consideration for interpreting our results is our non-population-representative snowball sampling strategy. Participants were volunteers recruited primarily from social networks of community organizers with our partner community organization and might differ in several ways from the eastern North Carolina population in general. Another consideration is our enrollment period from February 2021 to July 2022, including changing recommendations on vaccination and boosters, as well as increasing cases due to the more contagious Delta and Omicron variants. Vaccination, exposure, and treatment options varied for participants over the course of enrollment and differences over time may obscure differences by study group or participant characteristics. The long enrollment period also complicates comparisons with other southern US cohorts and CDC nationwide studies (Figure S2). Our study also has limitations associated with antibody test characteristics. Our multiplex assay was optimized for specificity over sensitivity, so we may have missed a proportion of infection-induced and vaccination or infection-induced antibody responses among our participants, especially for those infected a longer time ago who may have sero-reverted. The half-life of SARS-CoV-2 N IgG using our assay was about 64 days, compared to RBD about 100 days (45). Because we sampled participants through July 18, 2022, 910 days after the first confirmed case in the US, we likely underestimated infection-induced SARS-CoV-2 exposure.

Our findings show high rates of SARS-CoV-2 infection-induced IgG in a predominantly rural, Black, and Hispanic/Latino North Carolina cohort, especially among industrial livestock operation workers and their families. We add to reports of high numbers of cases associated with meatpacking facilities early in the course of the pandemic and to evidence of health disparities in exposure to SARS-CoV-2 by socioeconomic position. Delays in the timing of receipt of COVID-19 vaccination reinforce the importance of dismantling vaccination barriers, especially for industrial livestock operation workers and their household members. Associations between masking and physical distancing with antibody results also add to evidence of the effectiveness of these prevention strategies.

## MATERIALS AND METHODS

### Study design and participants

This study was designed and conducted in partnership with REACH. Data were collected by REACH community organizers and researchers from Johns Hopkins Bloomberg School of Public Health (JHSPH). Using a snowball sampling approach, REACH community organizers recruited ILO, ILON, and metro-area households. All enrolled households had at least one adult (≥18 years old) enrolled into the study. In addition to the one adult, all additional household members of any age were eligible to be enrolled. Eligibility criteria for all groups also included ability to understand spoken English or Spanish and access to household phone or mobile device and refrigerator. The study was developed in collaboration between JHSPH and REACH. The JHSPH Institutional Review Board (IRB) approved this study (IRB00014420).

### Questionnaire data and saliva sample collection

Before participation, adult participants provided oral consent. For children 0-6 years old, a parent or legal guardian provided permission, oral assent, and questionnaire responses for the child. For children 7-17 years old, a parent/legal guardian provided permission for the child and the child provided oral assent and questionnaire responses, with parents answering some questions as appropriate (e.g., health history). Recruitment, consent, questionnaires, and saliva self-collection were conducted remotely via video or phone call, without physical contact between study team and participants. Questionnaire responses were recorded and training and supervision of biospecimen self-collection were provided during the same video or phone call. Participants and parents or legal guardians of children 0-6 reported demographic information; work, school, or childcare outside the home; infection prevention behaviors; and health history, including information related to COVID-19 vaccination and symptoms consistent with COVID-19. Participants who worked at an ILO were also asked more detailed questions about livestock production and processing activities. Study questionnaires were developed in collaboration with REACH organizers. REACH interviewers included those fluent in English and Spanish, and participants had the ability to respond in either language. REACH and JHSPH interviewers recorded participant responses in REDCap, a secure web application for managing online surveys (48, 49).

After consent, all enrolled households received a study package containing all materials for saliva self-collection, self-collection procedure information, and packaging materials via REACH drop-off or direct shipping. During the questionnaire, training, and sampling call, REACH or JHSPH interviewers instructed all enrolled participants on how to collect saliva samples and stayed on the call as participants collected samples to answer any questions and ascertain if participants followed procedures. All participants provided two self-collected saliva samples: an oral fluid saliva sample and a passive drool saliva sample. For the oral fluid sample, participants brushed the Oracol+ 2.0 saliva collection device (Malvern Medical Developments, Worcester, United Kingdom) along their gums for 1-2 minutes. Participants were instructed to store their samples in a refrigerator until pickup by a REACH courier or direct shipping to JHSPH. Because booster vaccination was recommended by the CDC during the course of our study, we added questions about this topic mid-study and recontacted participants not initially asked.

### Multiplex SARS-CoV-2 IgG assay for oral fluid

Oral fluid samples were separated from sponges by centrifugation and tested for SARS-CoV-2 nucleocapsid (N), receptor-binding domain (RBD), and spike (S) IgG, using a multiplex bead-based immunoassay based on Luminex technology, which has been described previously (N. Pisanic, A. Antar, K. Kruczynski, M. G. Rivera, K. Spicer, P. R. Randad, A. Pekosz, S. L. Klein, M. J. Betenbaugh, B. Detrick, W. Clarke, D. L. Thomas, Y. C. Manabe, and C. D. Heaney, submitted for publication) (19, 20). Briefly, the multiplex assay included SARS-CoV-2 N, RBD, and S antigens coupled to magnetic microparticles, and a background control bead coated with bovine serum albumin (BSA). Saliva samples were centrifuged for 5 min at 10,000 g and 10 μL supernatant were added to a 96-well assay plate containing 40 μL bead mix (1,000 beads per bead set) in assay buffer (PBS-TBN). After incubation to allow for binding of SARS-CoV-2 specific IgG present in saliva samples, beads were washed, and fluorophore-labeled anti-human IgG was added to the plate. After a second incubation to allow for binding of labeled anti-IgG to salivary IgG on the beads, the plate was washed again, and median fluorescent intensity (MFI) was read on a Luminex MagPix instrument.

To determine optimum performance cutoffs for infection-induced IgG, we used 1320 saliva samples from individuals without known prior exposure to the SARS-CoV-2 virus or vaccine (presumed negatives) and 325 saliva samples collected >14 days after symptom onset of a molecularly confirmed SARS-CoV-2 infection (infection-induced positives). To determine optimum performance cutoffs for infection- and/or vaccination-induced IgG, we used 1002 saliva samples from individuals without known prior exposure to the SARS-CoV-2 virus or vaccine (presumed negatives) and 492 saliva samples collected >14 days after symptom onset of a molecularly confirmed SARS-CoV-2 infection and/or >14 days after completing the primary COVID-19 vaccination series (infection- and/or vaccination-induced positives). We first subtracted the BSA signal from all SARS-CoV-2 signals. The best algorithm for infection-induced IgG relied on N (Cat. No. Z03480, Genscript, NJ, USA) and RBD (Cat. No. 40592-V08H, Sino Biological, Beijing, China) (sensitivity=97.6%, specificity=99.4%) and the best algorithm for infection- and/or vaccination-induced IgG response relied on RBD (Cat. No. 40592-V08H, Sino Biological, Beijing, China) (sensitivity=99.4%, specificity=99.3%).

### Data from other southern US cohorts

Seropositivity data from the COVID-19 Prevention in Emory Healthcare Personnel (COPE) Study cohort were obtained through correspondence (M. H. Collins and C. D. Heaney, correspondence). Participants were health care providers recruited from 4 university-affiliated hospitals and clinics in Atlanta, Georgia, and saliva samples for serology were collected at enrollment, at 3 months, and at 6 months. Oral fluid saliva samples were tested with the multiplex assay described above, as in our North Carolina study population (19, 20, 29, 30). Monthly seropositivity and N seropositivity data from the Cabarrus County COVID-19 Prevalence and Immunity (C3PI) were also obtained through correspondence (L. K. Newby and D. Wixted, correspondence). Participants were selected from a larger ongoing cohort study through a weighted, randomized scheme to approximate the sex, age, and race/ethnicity of Cabarrus County, North Carolina. Blood samples for serology were collected monthly, and serology testing was performed with the Abbott Alinity IgG N protein antibody assay (specificity 99.9% and sensitivity 100%) (31). Nationwide and North Carolina infection-induced antibody seroprevalence estimates from CDC commercial laboratory surveys were downloaded from https://covid.cdc.gov/covid-data-tracker/#national-lab (36).

### Statistical analysis

We first compared the distribution of demographic characteristics and potential risk factors for SARS-CoV-2 IgG among the ILO household, ILON household, and metropolitan area (Metro) household groups. Using participants’ reported dates of receiving a first dose, receiving a second dose (or first dose for the Janssen [Johnson & Johnson]), and receiving a booster dose of the COVID-19 vaccine, we plotted time to each vaccination event by group and tested for difference in time to each vaccination event by group using the 3-group log-rank test implemented in *survdiff* function (*survival* package) in R. Next, we calculated the crude prevalence of salivary SARS-CoV-2 IgG outcomes in each household group (ILO, ILON, and Metro), and the crude prevalence of infection-induced IgG across levels of participant characteristics. For SARS-CoV-2 IgG and self-reported COVID-19 outcomes, we used generalized estimating equation (GEE) log-binomial regression models clustered by household to calculate crude prevalence ratios (PRs) and 95% confidence intervals (CIs) comparing outcome prevalence among the ILO versus the ILON, Metro, and combined ILON and Metro groups. We also used GEE log-binomial regression models to calculate crude prevalence ratios of infection-induced IgG by participant characteristics, with the category with the greatest number of participants as the reference group. To compare infection-induced IgG prevalence in our cohort to other southern US cohorts, we used log-binomial regression models to calculate crude prevalence ratios between the ILO group versus the ILON, Metro, and other southern US cohort groups with enrollment dates overlapping at least one day of our enrollment date range. All statistical analyses were completed in R 2022.02.0 (50).

## Supporting information

STROBE Checklist

## Data Availability

Data produced in the present study are available upon reasonable request to the authors.

## ACKNOWLEDGEMENTS

This study was supported by an anonymous gift, the JHU COVID-19 Research and Response Program, the FIA Foundation, and NIAID R21 R21AI139784. C.G., K.M.K., K. Koehler, and C.D.H. were supported by NIOSH ERC T42 OH0008428. Additionally, N.P., K. Kruczynski, M.G.R., K.S., and C.D.H. were supported by an anonymous gift, the JHU COVID-19 Research and Response Program, and the FIA Foundation. The C3PI Study was funded by research grants to Duke University from the NCDHHS and the Center for Disease Control and Prevention (CDC). The MURDOCK Study was funded by a gift from the David H. Murdock Institute for Business and Culture and is supported by Duke’s NIH National Center for Advancing Translational Sciences (NCATS) Clinical and Translational Science Award (CTSA) UL1TR002553. Additional support was provided by the Duke Claude D. Pepper Older Americans Independence Center Grant, 5P30AG028716-15.

We thank the community organizers at the Rural Empowerment Association for Community Help (REACH), especially Margaret Carr, Clesha Hall, Unique Hall, Angela Matthews, Arika Miller, and Helen Santizo, as well as all participants, without whom this study would not have been possible. We acknowledge Caryn Kok and Eric Xu for assistance with antibody testing. We also acknowledge Dr. Matthew H. Collins for sharing antibody prevalence data from the COVID-19 Prevention in Emory Healthcare Personnel (COPE) Study and Douglas Wixted and Dr. L. Kristin Newby for sharing antibody prevalence data from the C3PI Study.

## Notes

### Competing Interest Statement

The authors have declared no competing interest.

### Funding Statement

This study was funded by an anonymous gift, the JHU COVID-19 Research and Response Program, the FIA Foundation, and NIAID R21 R21AI139784. C.G., K.M.K., K. Koehler, and C.D.H. were supported by NIOSH ERC T42 OH0008428. Additionally, N.P., K. Kruczynski, M.G.R, K.S., and C.D.H were supported by an anonymous gift, the JHU COVID-19 Research and Response Program, and the FIA Foundation.

### Author Declarations

The Institutional Review Board (IRB) of Johns Hopkins Bloomberg School of Public Health gave ethical approval for this work (IRB00014420).

## REFERENCES

1. USDA. 2022. Annual Statistical Bulletin. USDA National Agricultural Statistics Service, North Carolina Field Office. https://www.nass.usda.gov/Statistics_by_State/North_Carolina/Publications/Annual_Statistical_Bulletin/index.php. Retrieved 12 April 2022.

2. Azaroff LS, Levenstein C, Wegman DH. 2002. Occupational injury and illness surveillance: conceptual filters explain underreporting. Am J Public Health 92:1421–1429.

3. Leigh JP, D. J, McCurdy SA. 2014. An estimate of the U.S. government’s undercount of nonfatal occupational injuries and illnesses in agriculture. Annals of Epidemiology 24:254–259.

4. Ceryes CA, Heaney CD. 2019. “Ag-gag” laws: evolution, resurgence, and public health implications. New Solutions: A Journal of Environmental and Occupational Health Policy 28:664–682.

5. BLS. Incidence rates of nonfatal occupational injuries and illnesses by industry and case types, 2020. US Bureau of Labor Statistics: Injuries, Illnesses, and Fatalities. https://www.bls.gov/iif/oshwc/osh/os/summ1_00_2020.htm. Retrieved 5 October 2022.

6. Donham KJ. 2010. Community and occupational health concerns in pork production: a review. Journal of Animal Science 88:E102–E111.

7. Wing S, Wolf S. 2000. Intensive livestock operations, health, and quality of life among eastern North Carolina residents. Environmental Health Perspectives 108:6.

8. Cole D, Todd L, Wing S. 2000. Concentrated swine feeding operations and public health: a review of occupational and community health effects. Environ Health Perspect 108:685–699.

9. Dyal JW, Grant MP, Broadwater K, Bjork A, Waltenburg MA, Gibbins JD, Hale C, Silver M, Fischer M, Steinberg J, Basler CA, Jacobs JR, Kennedy ED, Tomasi S, Trout D, Hornsby-Myers J, Oussayef NL, Delaney LJ, Patel K, Shetty V, Kline KE, Schroeder B, Herlihy RK, House J, Jervis R, Clayton JL, Ortbahn D, Austin C, Berl E, Moore Z, Buss BF, Stover D, Westergaard R, Pray I, DeBolt M, Person A, Gabel J, Kittle TS, Hendren P, Rhea C, Holsinger C, Dunn J, Turabelidze G, Ahmed FS, deFijter S, Pedati CS, Rattay K, Smith EE, Luna-Pinto C, Cooley LA, Saydah S, Preacely ND, Maddox RA, Lundeen E, Goodwin B, Karpathy SE, Griffing S, Jenkins MM, Lowry G, Schwarz RD, Yoder J, Peacock G, Walke HT, Rose DA, Honein MA. 2020. COVID-19 among workers in meat and poultry processing facilities — 19 states, April 2020. MMWR Morb Mortal Wkly Rep 69.

10. Waltenburg MA, Victoroff T, Rose CE, Butterfield M, Jervis R, Fedak K, Gabel JA, Feldpausch A, Dunne EM, Austen C, Ahmed FS, Tubach S, Rhea C, Krueger A, Crum DA, Vostok J, Moore MJ, Turabelidze G, Stover D, Donahue M, Edge K, Gutierrez B, Kline KE, Martz N, Cummins J, Barbeau B, Murphy J, Darby B, Graff NR, Dostal TKH, Pray IW, Tillman C, Dittrich MM, Burns-Grant G, Lee S, Spieckerman A, Iqbal K, Griffing SM, Lawson A, Mainzer HM, Bealle AE, Edding E, Arnold KE, Rodriguez T, Merkle S, Pettrone K, Schlanger K, LaBar K, Hendricks K, Lasry A, Krishnasamy V, Walke HT, Rose DA, Honein MA, COVID-19 Response Team. 2020. Update: COVID-19 among workers in meat and poultry processing facilities — United States, April–May 2020. MMWR Morb Mortal Wkly Rep 69:887–892.

11. 2021. House Select Subcommittee on the Coronavirus Crisis: How The Meatpacking Industry Failed the Workers Who Feed America. Staff Memorandum. Washington, DC. https://coronavirus.house.gov/sites/democrats.coronavirus.house.gov/files/2021.10.27%20Meatpacking%20Report.Final_.pdf.

12. Taylor CA, Boulos C, Almond D. 2020. Livestock plants and COVID-19 transmission. Proc Natl Acad Sci U S A 117:31706–31715.

13. Waltenburg MA, Rose CE, Victoroff T, Butterfield M, Dillaha JA, Heinzerling A, Chuey M, Fierro M, Jervis RH, Fedak KM, Leapley A, Gabel JA, Feldpausch A, Dunne EM, Austin C, Pedati CS, Ahmed FS, Tubach S, Rhea C, Tonzel J, Krueger A, Crum DA, Vostok J, Moore MJ, Kempher H, Scheftel J, Turabelidze G, Stover D, Donahue M, Thomas D, Edge K, Gutierrez B, Berl E, McLafferty M, Kline KE, Martz N, Rajotte JC, Julian E, Diedhiou A, Radcliffe R, Clayton JL, Ortbahn D, Cummins J, Barbeau B, Carpenter S, Pringle JC, Murphy J, Darby B, Graff NR, Dostal TKH, Pray IW, Tillman C, Rose DA, Honein MA. 2021. Coronavirus Disease among Workers in Food Processing, Food Manufacturing, and Agriculture Workplaces. Emerging Infectious Diseases 27:243–249.

14. Tai DBG, Shah A, Doubeni CA, Sia IG, Wieland ML. 2021. The Disproportionate Impact of COVID-19 on Racial and Ethnic Minorities in the United States. Clinical Infectious Diseases 72:703–706.

15. Zalla LC, Martin CL, Edwards JK, Gartner DR, Noppert GA. 2021. A Geography of Risk: Structural Racism and COVID-19 Mortality in the United States. American Journal of Epidemiology 190:1439–1446.

16. Lopez CA, Cunningham CH, Pugh S, Brandt K, Vanna UP, Delacruz MJ, Guerra Q, Bhowmik DR, Goldstein SJ, Hou YJ, Gearhart M, Wiethorn C, Pope C, Amditis C, Pruitt K, Newberry-Dillon C, Schmitz JL, Premkumar L, Adimora AA, Baric RS, Emch M, Boyce RM, Aiello AE, Fosdick BK, Larremore DB, de Silva AM, Juliano JJ, Markmann AJ. 2022. Ethnoracial Disparities in SARS-CoV-2 Seroprevalence in a Large Cohort of Individuals in Central North Carolina from April to December 2020. mSphere 7:e0084121.

17. CDC. 2020. Cases, Data, and Surveillance. Centers for Disease Control and Prevention. https://www.cdc.gov/coronavirus/2019-ncov/covid-data/investigations-discovery/hospitalization-death-by-race-ethnicity.html. Retrieved 11 March 2021.

18. Sherman AC, Smith T, Zhu Y, Taibl K, Howard-Anderson J, Landay T, Pisanic N, Kleinhenz J, Simon TW, Espinoza D, Edupuganti N, Hammond S, Rouphael N, Shen H, Fairley JK, Edupuganti S, Cardona-Ospina JA, Rodriguez-Morales AJ, Premkumar L, Wrammert J, Tarleton R, Fridkin S, Heaney CD, Scherer EM, Collins MH. 2021. Application of SARS-CoV-2 Serology to Address Public Health Priorities. Front Public Health 9:744535.

19. Heaney CD, Pisanic N, Randad PR, Kruczynski K, Zhu X, Littlefield K, Patel EU, Shrestha R, Shoham S, Sullivan D, Gebo K, Hanley D, Quinn TC, Casadevall A, Zenilman JM, Pekosz A, Bloch EM, Tobian AAR. Comparative performance of multiplex salivary and commercially available serologic assays to detect SARS-CoV-2 IgG and neutralization titers 25.

20. Pisanic N, Randad PR, Kruczynski K, Manabe YC, Thomas DL, Pekosz A, Klein SL, Betenbaugh MJ, Clarke WA, Laeyendecker O, Caturegli PP, Larman HB, Detrick B, Fairley JK, Sherman AC, Rouphael N, Edupuganti S, Granger DA, Granger SW, Collins MH, Heaney CD. 2020. COVID-19 Serology at Population Scale: SARS-CoV-2-Specific Antibody Responses in Saliva. J Clin Microbiol 59.

21. Cervia C, Nilsson J, Zurbuchen Y, Valaperti A, Schreiner J, Wolfensberger A, Raeber ME, Adamo S, Weigang S, Emmenegger M, Hasler S, Bosshard PP, De Cecco E, Bächli E, Rudiger A, Stüssi-Helbling M, Huber LC, Zinkernagel AS, Schaer DJ, Aguzzi A, Kochs G, Held U, Probst-Müller E, Rampini SK, Boyman O. 2021. Systemic and mucosal antibody responses specific to SARS-CoV-2 during mild versus severe COVID-19. J Allergy Clin Immunol 147:545-557.e9.

22. Russell MW, Moldoveanu Z, Ogra PL, Mestecky J. 2020. Mucosal Immunity in COVID-19: A Neglected but Critical Aspect of SARS-CoV-2 Infection. Frontiers in Immunology 11:3221.

23. Duarte N, Yanes-Lane M, Arora RK, Bobrovitz N, Liu M, Bego MG, Yan T, Cao C, Gurry C, Hankins CA, Cheng MP, Gingras A-C, Mazer BD, Papenburg J, Langlois M-A. 2022. Adapting Serosurveys for the SARS-CoV-2 Vaccine Era. Open Forum Infectious Diseases 9:ofab632.

24. Jones JM, Opsomer JD, Stone M, Benoit T, Ferg RA, Stramer SL, Busch MP. 2022. Updated US Infection- and Vaccine-Induced SARS-CoV-2 Seroprevalence Estimates Based on Blood Donations, July 2020-December 2021. JAMA 328:298–301.

25. CDC. 2022. CDC Museum COVID-19 Timeline. Centers for Disease Control and Prevention. https://www.cdc.gov/museum/timeline/covid19.html. Retrieved 1 September 2022.

26. CDC. 2022. Coronavirus Disease 2019 (COVID-19) – Symptoms. Centers for Disease Control and Prevention. https://www.cdc.gov/coronavirus/2019-ncov/symptoms-testing/symptoms.html. Retrieved 10 August 2022.

27. CDC. 2022. Weekly U.S. Influenza Surveillance Report. Centers for Disease Control and Prevention. https://www.cdc.gov/flu/weekly/index.htm. Retrieved 12 August 2022.

28. CDC. 2022. People with Certain Medical Conditions. Centers for Disease Control and Prevention. https://www.cdc.gov/coronavirus/2019-ncov/need-extra-precautions/people-with-medical-conditions.html. Retrieved 12 August 2022.

29. Howard-Anderson JR, Adams C, Dube WC, Smith TC, Sherman AC, Edupuganti N, Mendez M, Chea N, Magill SS, Espinoza DO, Zhu Y, Phadke VK, Edupuganti S, Steinberg JP, Lopman BA, Jacob JT, Fridkin SK, Collins MH. 2022. Occupational risk factors for severe acute respiratory coronavirus virus 2 (SARS-CoV-2) infection among healthcare personnel: A 6-month prospective analysis of the COVID-19 Prevention in Emory Healthcare Personnel (COPE) Study. Infect Control Hosp Epidemiol Online ahead of print:1–8.

30. Howard-Anderson JR, Adams C, Sherman AC, Dube WC, Smith TC, Edupuganti N, Chea N, Magill SS, Espinoza DO, Zhu Y, Phadke VK, Edupuganti S, Steinberg JP, Lopman BA, Jacob JT, Collins MH, Fridkin SK. 2022. Occupational risk factors for severe acute respiratory coronavirus virus 2 (SARS-CoV-2) infection among healthcare personnel: A cross-sectional analysis of subjects enrolled in the COVID-19 Prevention in Emory Healthcare Personnel (COPE) study. Infect Control Hosp Epidemiol 43:381–386.

31. Neighbors CE, Wu AE, Wixted DG, Heidenfelder BL, Kingsbury CA, Register HM, Louzao R, Sloane R, Eckstrand J, Pieper CC, Faldowski RA, Denny TN, Woods CW, Newby LK. 2022. The Cabarrus County COVID-19 Prevalence and Immunity (C3PI) Study: design, methods, and baseline characteristics. Am J Transl Res 14:5693–5711.

32. Saitone TL, Aleks Schaefer K, Scheitrum DP. 2021. COVID-19 morbidity and mortality in U.S. meatpacking counties. Food Policy 101:102072.

33. Mitloehner FM, Calvo MS. 2008. Worker health and safety in concentrated animal feeding operations. J Agric Saf Health 14:163–187.

34. Campbell DS. 1999. Health hazards in the meatpacking industry. Occup Med 14:351–372.

35. Ma Q, Liu J, Liu Q, Kang L, Liu R, Jing W, Wu Y, Liu M. 2021. Global Percentage of Asymptomatic SARS-CoV-2 Infections Among the Tested Population and Individuals With Confirmed COVID-19 Diagnosis: A Systematic Review and Meta-analysis. JAMA Network Open 4:e2137257.

36. CDC. 2020. COVID Data Tracker. Centers for Disease Control and Prevention. https://covid.cdc.gov/covid-data-tracker. Retrieved 7 June 2021.

37. Nguyen KH, Nguyen K, Geddes M, Allen JD, Corlin L. 2022. Trends in COVID-19 vaccination receipt and intention to vaccinate, United States, April to August, 2021. American Journal of Infection Control 50:699–703.

38. State-by-state data on COVID-19 vaccinations in the United States. Our World in Data. https://ourworldindata.org/us-states-vaccinations. Retrieved 23 August 2022.

39. Bajema KL, Wiegand RE, Cuffe K, Patel SV, Iachan R, Lim T, Lee A, Moyse D, Havers FP, Harding L, Fry AM, Hall AJ, Martin K, Biel M, Deng Y, Meyer WA, Mathur M, Kyle T, Gundlapalli AV, Thornburg NJ, Petersen LR, Edens C. 2021. Estimated SARS-CoV-2 Seroprevalence in the US as of September 2020. JAMA Intern Med 181:450–460.

40. Howard J, Huang A, Li Z, Tufekci Z, Zdimal V, van der Westhuizen H-M, von Delft A, Price A, Fridman L, Tang L-H, Tang V, Watson GL, Bax CE, Shaikh R, Questier F, Hernandez D, Chu LF, Ramirez CM, Rimoin AW. 2021. An evidence review of face masks against COVID-19. Proceedings of the National Academy of Sciences 118:e2014564118.

41. Pathak EB, Menard J, Garcia RB, Salemi JL. 2021. Social Class, Race/Ethnicity, and COVID-19 Mortality Among Working Age Adults in the United States.

42. Pathak EB, Menard JM, Garcia RB, Salemi JL. 2022. Joint Effects of Socioeconomic Position, Race/Ethnicity, and Gender on COVID-19 Mortality among Working-Age Adults in the United States. Int J Environ Res Public Health 19:5479.

43. Paul A. Landsbergis, Joseph G. Grzywacz, Anthony D. LaMontagne. 2014. Work organization, job insecurity, and occupational health disparities. American Journal of Industrial Medicine 57:495–515.

44. Chu DK, Akl EA, Duda S, Solo K, Yaacoub S, Schünemann HJ, Chu DK, Akl EA, El-harakeh A, Bognanni A, Lotfi T, Loeb M, Hajizadeh A, Bak A, Izcovich A, Cuello-Garcia CA, Chen C, Harris DJ, Borowiack E, Chamseddine F, Schünemann F, Morgano GP, Schünemann GEUM, Chen G, Zhao H, Neumann I, Chan J, Khabsa J, Hneiny L, Harrison L, Smith M, Rizk N, Rossi PG, AbiHanna P, El-khoury R, Stalteri R, Baldeh T, Piggott T, Zhang Y, Saad Z, Khamis A, Reinap M, Duda S, Solo K, Yaacoub S, Schünemann HJ. 2020. Physical distancing, face masks, and eye protection to prevent person-to-person transmission of SARS-CoV-2 and COVID-19: a systematic review and meta-analysis. The Lancet 395:1973–1987.

45. Randad PR, Pisanic N, Kruczynski K, Howard T, Rivera MG, Spicer K, Antar AAR, Penson T, Thomas DL, Pekosz A, Ndahiro N, Aliyu L, Betenbaugh MJ, Manley H, Detrick B, Katz M, Cosgrove S, Rock C, Zyskind I, Silverberg JI, Rosenberg AZ, Duggal P, Manabe YC, Collins MH, Heaney CD. 2021. Durability of SARS-CoV-2-specific IgG responses in saliva for up to 8 months after infection.

46. Mutambudzi M, Niedzwiedz CL, Macdonald EB, Leyland AH, Mair FS, Anderson JJ, Celis-Morales CA, Cleland J, Forbes J, Gill JM, Hastie C, Ho FK, Jani BD, Mackay DF, Nicholl BI, O’Donnell CA, Sattar NI, Welsh PI, Pell JP, Katikireddi SV, Demou E. 2020. Occupation and risk of COVID-19: prospective cohort study of 120,621 UK Biobank participants. medRxiv 2020.05.22.20109892.

47. Nguyen LH, Drew DA, Graham MS, Joshi AD, Guo C-G, Ma W, Mehta RS, Warner ET, Sikavi DR, Lo C-H, Kwon S, Song M, Mucci LA, Stampfer MJ, Willett WC, Eliassen AH, Hart JE, Chavarro JE, Rich-Edwards JW, Davies R, Capdevila J, Lee KA, Lochlainn MN, Varsavsky T, Sudre CH, Cardoso MJ, Wolf J, Spector TD, Ourselin S, Steves CJ, Chan AT, Albert CM, Andreotti G, Bala B, Balasubramanian BA, Beane-Freeman LE, Brownstein JS, Bruinsma FJ, Coresh J, Costa R, Cowan AN, Deka A, Deming-Halverson SL, Elena Martinez M, Ernst ME, Figueiredo JC, Fortuna P, Franks PW, Freeman LB, Gardner CD, Ghobrial IM, Haiman CA, Hall JE, Kang JH, Kirpach B, Koenen KC, Kubzansky LD, Lacey, Jr JV, Le Marchand L, Lin X, Lutsey P, Marinac CR, Martinez ME, Milne RL, Murray AM, Nash D, Palmer JR, Patel AV, Pierce E, Robertson MM, Rosenberg L, Sandler DP, Schurman SH, Sewalk K, Sharma SV, Sidey-Gibbons CJ, Slevin L, Smoller JW., Steves CJ, Tiirikainen MI, Weiss ST, Wilkens LR, Zhang F. 2020. Risk of COVID-19 among front-line health-care workers and the general community: a prospective cohort study. The Lancet Public Health S246826672030164X.

48. Harris PA, Taylor R, Thielke R, Payne J, Gonzalez N, Conde JG. 2009. Research electronic data capture (REDCap)--a metadata-driven methodology and workflow process for providing translational research informatics support. J Biomed Inform 42:377–381.

49. Harris PA, Taylor R, Minor BL, Elliott V, Fernandez M, O’Neal L, McLeod L, Delacqua G, Delacqua F, Kirby J, Duda SN. 2019. The REDCap Consortium: Building an International Community of Software Platform Partners. J Biomed Inform 95:103208.

50. R: The R Project for Statistical Computing. https://www.r-project.org/. Retrieved 18 October 2021.

