## Supplemental Information for "SARS-CoV-2 antibody prevalence among industrial livestock operation workers and nearby community residents, North Carolina, USA, 2021-2022"

***Table S1.*** *COVID-19 vaccination by study group compared to North Carolina and US by quartile of interview or follow-up call dates*

| Characteristic | ILO (n=90) | ILON (n=97) | Metro (n=92) | JHU-REACH study population (n=279) | North Carolina[a] | US[a] |
|---|---|---|---|---|---|---|
| Completed initial COVID-19 vaccination protocol, n (%) | | | | | | |
| By December 29, 2021 (Q1) | 6/19 (31.6) | 18/35 (51.4) | 8/16 (50) | 32/70 (45.7) | (56.7) | (61.9) |
| By January 10, 2022 (Q2) | 12/33 (36.4) | 40/70 (57.1) | 19/37 (51.3) | 71/140 (50.7) | (57.3) | (62.6) |
| By March 3, 2022 (Q3) | 32/60 (53.3) | 44/96 (45.8) | 28/54 (51.9) | 105/210 (50) | (59.4) | (65) |
| By July 18, 2022 (Q4) | 47 (51.6) | 47 (48.5) | 51 (55.4) | 145 (52) | (62.9) | (67.2) |
| Received at least one booster dose, n (%) | | | | | | |
| By December 29, 2021 (Q1) | 1/19 (5.3) | 1/35 (2.9) | 0 (0) | 2/70 (2.9) | (11.3) | (20.5) |
| By January 10, 2022 (Q2) | 3/33 (9.1) | 9/70 (12.9) | 5/37 (13.5) | 17/140 (12.1) | (12.5) | (22.8) |
| By March 3, 2022 (Q3) | 6/60 (10) | 10/96 (10.4) | 10/54 (18.5) | 26/210 (12.4) | (15) | (28.5) |
| By July 18, 2022 (Q4) | 12 (13.2) | 10 (10.3) | 19 (20.7) | 41 (14.7) | (17.7) | (32.3) |
| Answered follow-up call, n (%) | 73 (81.1) | 88 (90.7) | 82 (89.1) | 243 (87.1) | - | - |

| | | | | | | |
|---|---|---|---|---|---|---|
| Follow-up or interview (if did not answer follow-up) call dates, median (IQR) | 2/10/2022 (12/29/2021-3/29/2022) | 1/3/2022 (11/3/2021-1/11/2022) | 1/11/2022 (1/03/2022-4/11/2022) | 1/10/2022 (12/29/2021-3/2/2022) | - | - |

*[a]US CDC estimates from Our World in Data visualization (1)*

*Note: ILO refers to study participants living in a household with at least one adult working at an industrial hog or poultry operation, meatpacking plant, or animal rendering plant; ILON refers to participants living nearby these facilities without any known occupational exposure to livestock; Metro refers to participants living in metropolitan areas of North Carolina. Participants who reported vaccination but did not provide a corresponding date are included as by July 18, 2022, final date of interviews and follow-up calls.*

***Table S2.*** *Self-reported infection prevention behaviors and health history by study group, North Carolina, USA, 2021-2022*

| Characteristic, n (%) | ILO n=90 (80 households) | | ILON n=97 (80 households) | | Metro n=92 (80 households) | | p-value[a] |
|---|---|---|---|---|---|---|---|
| **Infection prevention behaviors** | | | | | | | |
| Reduced physical contact with people outside your home | | | | | | | 0.16 |
| Yes, all household members | 65 | (71.4) | 81 | (83.5) | 69 | (75) | |
| Yes, some but not all household members | 10 | (11) | 4 | (4.1) | 6 | (6.5) | |
| No | 9 | (9.9) | 4 | (4.1) | 7 | (7.6) | |
| No response | 7 | (7.7) | 8 | (8.2) | 10 | (10.9) | |
| Avoiding or cancelling travel or vacation plans | | | | | | | 0.05 |
| Yes | 72 | (79.1) | 75 | (77.3) | 56 | (60.9) | |
| No | 19 | (20.9) | 21 | (21.6) | 33 | (35.9) | |
| No response | 0 | | 1 | (1) | 3 | (3.3) | |
| Wearing a mask when out in public | | | | | | | 0.16 |
| Yes | 86 | (94.5) | 93 | (95.9) | 83 | (90.2) | |
| No | 5 | (5.5) | 3 | (3.1) | 9 | (9.8) | |

| | | | | | | | |
|---|---|---|---|---|---|---|---|
| No response | 0 | (0) | 1 | (1) | 0 | (0) | |
| Washing hands/using hand sanitizer more frequently | | | | | | | 0.62 |
| Yes | 88 | (96.7) | 90 | (92.8) | 86 | (93.5) | |
| No | 3 | (3.3) | 5 | (5.2) | 5 | (5.4) | |
| No response | 0 | (0) | 2 | (2.1) | 1 | (1.1) | |
| At times (last 2 w) you interacted with people inside … | | | | | | | |
| How often did you maintain a 6 ft distance from others? | | | | | | | 0.41 |
| Always | 67 | (73.6) | 78 | (80.4) | 68 | (73.9) | |
| Sometimes | 24 | (26.4) | 17 | (17.5) | 21 | (22.8) | |
| Never | 0 | (0) | 1 | (1) | 2 | (2.2) | |
| No response | 0 | (0) | 1 | (1) | 1 | (1.1) | |
| How often did you wear a mask? | | | | | | | 0.06 |
| Always | 74 | (81.3) | 82 | (84.5) | 66 | (71.7) | |
| Sometimes | 17 | (18.7) | 13 | (13.4) | 22 | (23.9) | |
| Never | 0 | (0) | 1 | (1) | 4 | (4.3) | |
| No response | 0 | (0) | 1 | (1) | 0 | (0) | |

| | | | | | | | |
|---|---|---|---|---|---|---|---|
| **Health history** | | | | | | | |
| Do you have any of the following chronic conditions? | | | | | | | |
| Hypertension | 25 | (27.5) | 28 | (28.9) | 22 | (23.9) | 0.77 |
| Diabetes | 15 | (16.5) | 17 | (17.5) | 10 | (10.9) | 0.43 |
| Asthma | 9 | (9.9) | 5 | (5.2) | 13 | (14.1) | 0.10 |
| Depression | 2 | (2.2) | 6 | (6.2) | 12 | (13) | 0.03 |
| Cardiovascular disease | 2 | (2.2) | 6 | (6.2) | 5 | (5.4) | 0.38 |
| Cancer diagnosis/treatment (past 12 mo) | 4 | (4.4) | 3 | (3.1) | 1 | (1.1) | 0.41 |
| Chronic kidney disease | 1 | (1.1) | 4 | (4.1) | 3 | 3.3 | 0.44 |
| Autoimmune disease | 1 | (1.1) | 2 | (2.1) | 2 | 2.2 | 0.83 |
| Chronic obstructive pulmonary disease (COPD) | 2 | (2.2) | 1 | (1) | 1 | (1.1) | 0.76 |
| Immunocompromised condition | 2 | (2.2) | 0 | (0) | 1 | (1.1) | 0.35 |
| Other chronic lung disease | 2 | (2.2) | 1 | (1) | 0 | (0) | 0.36 |
| Sickle cell anemia | 1 | (1.1) | 0 | (0) | 1 | (1.1) | 0.59 |
| Do you have seasonal allergies? | 42 | (46.2) | 49 | (50.5) | 57 | (62) | 0.04 |

| | | | | | | | |
|---|---|---|---|---|---|---|---|
| Have you gotten or plan to get a flu shot this year? | 36 | (39.6) | 48 | (49.5) | 44 | (47.8) | 0.30 |
| Fever with a cough at the same time or fever with a sore throat at the same time (past yr)?[b] | 19 | (20.9) | 15 | (15.5) | 17 | (18.5) | 0.33 |
| Since February 1, 2020, have you thought you had COVID-19? | 22 | (24.2) | 18 | (18.6) | 18 | (19.6) | 0.28 |

[a]*p-value associated with Chi-square test statistic*

[b]*CDC definition of influenza-like illness* (2)

*Note: ILO refers to study participants living in a household with at least one adult working at an industrial hog or poultry operation, meatpacking plant, or animal rendering plant; ILON refers to participants living nearby these facilities without any known occupational exposure to livestock; Metro refers to participants living in metropolitan areas of North Carolina.*

| | Group | Date quartile | Black, Hispanic, Other | HS+ or not | >1 person per bedroom or not | Physical distancing | Masking or not | Vaccinated |
|---|---|---|---|---|---|---|---|---|
| Group | 1 | 0.39 | 0.17 | 0.25 | 0.23 | 0.11 | 0.12 | 0.11 |
| Date quartile | NA | 1 | 0.18 | 0.14 | 0.18 | 0.23 | 0.14 | 0.42 |
| Black, Hispanic, Other | NA | NA | 1 | 0.21 | 0.12 | 0.10 | 0.10 | 0.11 |
| HS+ or not | NA | NA | NA | 1 | 0.09 | 0.01 | 0.03 | 0.13 |
| >1 person per bedroom or not | NA | NA | NA | NA | 1 | 0.06 | 0.09 | 0.02 |
| Physical distancing | NA | NA | NA | NA | NA | 1 | 0.05 | 0.04 |
| Masking or not | NA | NA | NA | NA | NA | NA | 1 | 0.03 |
| Vaccinated | NA | NA | NA | NA | NA | NA | NA | 1 |

***Figure S1.*** *Correlation matrix (Cramer's V) between factors associated with SARS-CoV-2 infection-induced IgG prevalence, categorized as in Table 3.*

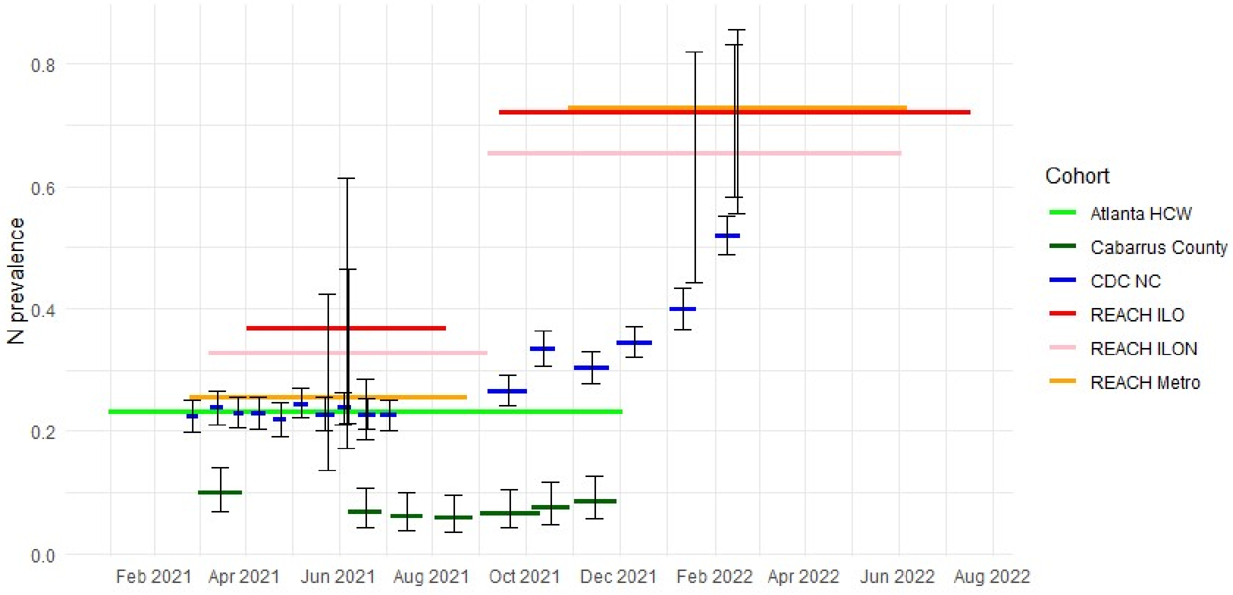


***Figure S2****. SARS-CoV-2 infection-induced IgG prevalence over time by cohort; REACH ILO, ILON, and Metro groups split into participants sampled before vs. after the median sampling date, September 6, 2021, for better comparability with other cohorts over time. Note: REACH refers to our study population and collaboration with the Rural Empowerment Association for Community Help (REACH); ILO refers to study participants living in a household with at least one adult working at an industrial hog or poultry operation, meatpacking plant, or animal rendering plant; ILON refers to participants living nearby these facilities without any known occupational exposure to livestock; Metro refers to participants living in metropolitan areas of North Carolina.*

***Table S3.*** *Selected participant characteristics by study group compared to Atlanta health care worker (HCW) cohort and Cabarrus County, NC general population cohort*

| **Characteristic, n (%)** | **ILO (n=90)** | **ILON (n=97)** | **Metro (n=92)** | **JHU-REACH study population (n=279)** | **Atlanta HCW (n=301)** | **Cabarrus County (n=300)** |
|---|---|---|---|---|---|---|
| SARS-CoV-2 infection-induced IgG[a] | 46/73 (63%) | 36/84 (43%) | 37/76 (49%) | 119/233 (51.1%) | 71/306 (23.2%) | 23/270 (8.5%) |
| Sampling date, median (range) | 2/9/2022 (4/2/2021-7/18/2022) | 6/6/2021 (3/8/2021-6/3/2022) | 8/18/2021 (2/23/2021-6/7/2022) | 9/6/2021 (2/23/2021-7/18/2021) | – | – |
| | | | | | Latest follow-up 6 months post-enrollment (1/2021-12/2021) | Latest follow-up 11/2021 |
| Age in years, median (range) | 41.5 (13-67) | 50 (5-83) | 37 (9-74) | 41 (5-83) | – | – |
| | | | | | 172 (57.1) <40 years | Median 56 (IQR 47, 67) |

| | | | | | | |
|---|---|---|---|---|---|---|
| Gender | | | | | | |
| Female | 47 (52) | 54 (56) | 67 (73) | 139 (59.7) | 231 (76.7) | 183 (61.0) |
| Male | 43 (48) | 42 (43) | 25 (27) | 94 (40.3) | 70 (23.3) | 117 (39.0) |
| No response | | | | | | |
| Race/ethnicity | | | | | | |
| Black/African American | 79 (88) | 83 (86) | 74 (80) | 236 (84.5) | 33 (11.0) | 51 (17.0) |
| Hispanic/Latino | 8 (9) | 11 (11.3) | 3 (3.3) | 22 (7.9) | 13 (4.3) | 33 (11.0) |
| White/Caucasian | 1 (1.1) | 1 (1) | 8 (8.7) | 10 (3.6) | 219 (72.8) | 230 (76.7) |
| Both Black and White | 0 (0) | 1 (1) | 2 (2.2) | 3 (1.1) | – | – |
| Asian-American | 0 (0) | 0 (0) | 1 (1) | 1 (0.004) | 29 (9.6) | 0 (0) |
| Other or no response | 2 (2.2) | 1 (1) | 3 (3.3) | 6 (2.2) | 20 (6.6) | – |
| Education | | | | | | |
| High school diploma/GED or less | 65 (71) | 53 (55) | 37 (40) | 155 (55.6) | – | 19 (6.3) |
| Post-high school | 26 (29) | 42 (43) | 53 (58) | 121 (43.3) | – | 180 (93.3) |
| No response | 0 (0) | 2 (2.2) | 2 (2.2) | 4 (1.4) | – | 1 (0.3) |

| | | | | | | |
|---|---|---|---|---|---|---|
| Household members, mean (SD) | 2.7 (1.4) | 2.1 (1.3) | 2.2 (1.2) | 2.3 (1.3) | – | Median 2 (IQR 2, 4) |
| Health insurance[b] | | | | | | |
| Company health insurance plan | 30 (33) | 33 (34) | 56 (61) | 119 (42.7) | – | 169 (56.3) |
| Public health insurance | 34 (38) | 35 (36) | 17 (19) | 86 (30.8) | – | 94 (31.3) |
| Private health insurance | 15 (17) | 16 (17) | 10 (11) | 41 (14.7) | – | 12 (4.2) |
| No health insurance | 12 (13) | 15 (16) | 8 (9) | 35 (12.5) | – | 14 (4.7) |
| Other or no response | 2 (2) | 0 (0) | 1 (1.1) | 3 (1.1) | – | 25 (8.3) |

[a]*Number of samples tested for SARS-CoV-2-specific N IgG does not necessarily match size of study population group. Not all JHU-REACH samples were tested; Atlanta HCW and Cabarrus County N IgG results are only shown for the latest follow-up visit; some Atlanta HCW participants from the latest follow-up visit were not included for analysis in the manuscript from which participant characteristics were extracted* (3)

[b]*Participants could select more than one option.*

*Note: ILO refers to study participants living in a household with at least one adult working at an industrial hog or poultry operation, meatpacking plant, or animal rendering plant; ILON refers to participants living nearby these facilities without any known occupational exposure to livestock; Metro refers to participants living in metropolitan areas of North Carolina; "—" refers to data not available.*

REFERENCES

1. State-by-state data on COVID-19 vaccinations in the United States. Our World in Data. https://ourworldindata.org/us-states-vaccinations. Retrieved 23 August 2022.

2. CDC. 2022. Weekly U.S. Influenza Surveillance Report. Centers for Disease Control and Prevention. https://www.cdc.gov/flu/weekly/index.htm. Retrieved 12 August 2022.

3. Howard-Anderson JR, Adams C, Dube WC, Smith TC, Sherman AC, Edupuganti N, Mendez M, Chea N, Magill SS, Espinoza DO, Zhu Y, Phadke VK, Edupuganti S, Steinberg JP, Lopman BA, Jacob JT, Fridkin SK, Collins MH. 2022. Occupational risk factors for severe acute respiratory coronavirus virus 2 (SARS-CoV-2) infection among healthcare personnel: A 6-month prospective analysis of the COVID-19 Prevention in Emory Healthcare Personnel (COPE) Study. Infect Control Hosp Epidemiol 1–8.
